# Characterizing Post-Acute Sequelae of SARS-CoV-2 Infection across Claims and Electronic Health Record Databases

**DOI:** 10.1101/2021.03.19.21253756

**Authors:** Matthew E. Spotnitz, George Hripcsak, Patrick B. Ryan, Karthik Natarajan

## Abstract

**Importance:** Post-acute sequelae of SARS-CoV-2 infection (PASC) is emerging as a major public health issue.

**Objective:** We characterized the incidence of PASC, or related symptoms and diagnoses, for COVID-19 and influenza patients.

**Design:** Retrospective cohort study.

**Setting:** Our data sources were the IBM MarketScan Commercial Claims and Encounters (CCAE), Optum Electronic Health Record (EHR) and Columbia University Irving Medical Center (CUIMC) databases that were transformed to the Observational Medical Outcome Partnership (OMOP) Common Data Model (CDM) and were part of the Observational Health Sciences and Informatics (OHDSI) network.

**Participants:** The COVID-19 cohort consisted of patients with a diagnosis of COVID-19 or positive lab test of SARS-CoV-2 after January 1st 2020 with a follow up period of at least 30 days. The influenza cohort consisted of patients with a diagnosis of influenza between October 1, 2018 and May 1, 2019 with a follow up period of at least 30 days.

**Intervention:** Infection with COVID-19 or influenza.

**Main Outcomes and Measures:** Post-acute sequelae of SARS-CoV-2 infection (PASC), or related diagnoses, for COVID-19 and influenza patients.

**Results:** In aggregate, we characterized the post-acute experience for over 440,000 patients who were diagnosed with COVID-19 or tested positive for SARS-COV-2. The long term sequelae that had a higher incidence in the COVID-19 compared to Influenza cohorts were altered smell or taste, myocarditis, acute kidney injury, dyspnea and alopecia. Additionally, the long term incidences of respiratory illness, musculoskeletal disease, and psychiatric disorders for the COVID-19 population were higher than expected.

**Conclusions and Relevance:** The long term sequelae of COVID-19 and influenza may be different. Further characterization of PASC on large scale observational healthcare databases is warranted.

## Background

The global pandemic of SARS-CoV-2 infection and impact of COVID-19 disease has resulted in major morbidity and mortality worldwide. While substantial research has sought to characterize the disease natural history and the acute management of COVID-19, comparatively fewer studies have focused on the post-acute sequelae of SARS-CoV-2 infection (PASC)^1-6^. Much of the publicly available data on PASC come from case reports and single-institution prospective cohort studies^7-13^. Patient reported symptom data have shown that prolonged fatigue, headache, dyspnea and anosmia are PASC symptoms^14^. However, there are fewer claims and electronic health record (EHR) data about the incidence of PASC symptoms. Therefore, the public knowledge about PASC symptom presentation is evolving. The National Institutes of Health has encouraged the study of PASC on large-scale observational databases in order to better understand the condition and its public health impact^15^. In alignment with that effort, we present data on PASC patients from EHR and claims databases with an aim to characterize the natural history of patients with SARS-CoV-2 who developed symptoms or diagnoses related to PASC. Additionally, we characterized related long-term sequelae of influenza to provide a comparison.

## Methods

Three observational health databases were used for the analysis: a private-payer administrative claims database (IBM MarketScan Commercial Claims and Encounters-CCAE), a database of inpatient and outpatient electronic health records (Optum© de-identified Electronic Health Record Dataset-Optum EHR), and an electronic health record system from an academic medical center (Columbia University Irving Medical Center-CUIMC). All databases were transformed to the Observational Medical Outcome Partnership (OMOP) Common Data Model (CDM), and described further in Supplementary Appendix#1.

We defined a COVID-19 cohort as patients with a diagnosis of COVID-19 or positive lab test of SARS-CoV-2 after January 1^st^ 2020 with a follow up period of at least 30 days. An influenza cohort was defined as patients with a diagnosis of influenza between October 1, 2018 and May 1, 2019 with a follow up period of at least 30 days. We performed characterizations of COVID-19 and influenza patients who had at least one of the following symptoms or diagnoses that were related to Post-Acute Sequelae of SARS-CoV-2 Infection (PASC), according to the Centers for Disease Control and Prevention (CDC)^15^: altered smell or taste, myocarditis, acute kidney injury, dyspnea, alopecia, tachycardia, chest pain, lung disorder, myalgias, dementia or cognitive Impairment, malaise, fatigue, stress disorder, depression, anxiety, joint pain, mood changes, cough, rash, and fever. We calculated the number of patients who had each and any of these diagnoses between 30 and 180 days after the index event.

Diagnoses were based on ICD-10-CM and SNOMED codes, while lab tests included LOINC codes. The full list of codes for the symptoms and diagnoses (Supplementary Appendix #2) and our calculation of relative risk (Supplementary Appendix#3) are provided in the supplementary information.

## Results

In aggregate, we characterized the post-acute experience for over 440,000 patients who were diagnosed with COVID-19 or tested positive for SARS-COV-2. We identified 119,510 patients with COVID-19 in the Optum EHR database. Of those, 42,991 (36.28%) had at least one long term sequela. In the IBM MarketScan CCAE database, we identified a total of 306,142 patients with COVID-19, 74,320 (24.28%) of whom had at least one long term sequela. In the CUIMC database, 6,198 (27.52%) of 22,524 patients of patients with COVID-19 had a PASC-related observation within the following 6 months.

Table 1 shows the number of patients from the COVID-19 and influenza cohorts in each database, and subgroups of patients with all or any PASC diagnoses. Five PASC diagnoses had higher relative risk in COVID-19 compared to Influenza patients: altered smell or taste, myocarditis, acute kidney injury, dyspnea and alopecia.

**Table 1:** The sizes of the COVID-19 and influenza cohorts for the Optum-EHR, IBM MarketScan CCAE and CUIMC 2020q4 databases are shown. The numbers and percentages of patients who had any or all post-acute sequelae of SARS-CoV-2 (PASC) infection diagnoses between 30 and 180 days after the index event are shown. Those subgroups are ranked in descending order of relative risk for COVID-19 compared to influenza patients. The red cells show data for the group of symptoms and conditions that had the highest weighted relative risk, yellow cells show data for the next to highest group, and green cells show the lowest group. N, number; %, percentage; EHR, Electronic Health Record; CCAE, Clinical Claims and Encounters; CUIMC, Columbia University Irving Medical Center; 2020q4, 2020 4^th^ Quarter; RR, Relative Risk.

|  | Optum EHR |  |  |  |  | IBM MarketScan CCAE |  |  |  |  | CUIMC - 2020Q4 |  |  |  |  | Weighted |
| --- | --- | --- | --- | --- | --- | --- | --- | --- | --- | --- | --- | --- | --- | --- | --- | --- |
|  | COVID-19 |  | influenza |  | RR | COVID-19 |  | influenza |  | RR | COVID-19 |  | influenza |  | RR | RR |
|  | N | % | N | % |  | N | % | N | % |  | N | % | N | % |  |  |
| Total | 119,510 |  | 232,815 |  |  | 306,142 |  | 568,873 |  |  | 22,524 |  | 2,182 |  |  |  |
| At least one long term sequela among below | 42,991 | 36.28% | 70,518 | 30.29% |  | 74,320 | 24.28% | 178,753 | 31.42% |  | 6,198 | 27.52% | 913 | 41.84% |  |  |
| Altered Smell or Taste | 460 | 0.39% | 71 | 0.03% | 13.00 | 913 | 0.30% | 193 | 0.03% | 10.00 | 91 | 0.40% | 2 | 0.09% | 4.44 | 10.90 |
| Myocarditis | 101 | 0.09% | 36 | 0.02% | 4.50 | 116 | 0.04% | 50 | 0.01% | 4.00 | 39 | 0.17% | 1 | 0.05% | 3.40 | 4.20 |
| Acute Kidney Injury | 2,778 | 2.34% | 1676 | 0.72% | 3.25 | 1,322 | 0.43% | 1,015 | 0.18% | 2.39 | 235 | 1.04% | 91 | 4.17% | 0.25 | 2.48 |
| Dyspnea | 8,127 | 6.86% | 5,970 | 2.56% | 2.68 | 12,584 | 4.11% | 10,765 | 1.89% | 2.17 | 1,250 | 5.55% | 173 | 7.93% | 0.70 | 2.28 |
| Alopecia | 905 | 0.76% | 667 | 0.29% | 2.62 | 1,905 | 0.62% | 2,222 | 0.39% | 1.59 | 150 | 0.67% | 5 | 0.23% | 2.91 | 2.08 |
| Tachycardia | 3,963 | 3.34% | 4003 | 1.72% | 1.94 | 6,277 | 2.05% | 8,519 | 1.50% | 1.37 | 593 | 2.63% | 74 | 3.39% | 0.78 | 1.57 |
| Chest Pain | 7,132 | 6.02% | 7,465 | 3.21% | 1.88 | 10,997 | 3.59% | 15,302 | 2.69% | 1.33 | 949 | 4.21% | 134 | 6.14% | 0.69 | 1.52 |
| Lung Disorder | 10,226 | 8.63% | 11,501 | 4.94% | 1.75 | 9,212 | 3.01% | 13,009 | 2.29% | 1.31 | 883 | 3.92% | 284 | 13.02% | 0.30 | 1.40 |
| Myalgias | 1,400 | 1.18% | 1630 | 0.70% | 1.69 | 2,874 | 0.94% | 5,293 | 0.93% | 1.01 | 185 | 0.82% | 14 | 0.64% | 1.28 | 1.30 |
| Dementia or Cognitive Impairment | 2,813 | 2.37% | 2277 | 0.98% | 2.42 | 1,148 | 0.37% | 2,515 | 0.44% | 0.84 | 256 | 1.14% | 56 | 2.57% | 0.44 | 1.35 |
| Malaise or Fatigue | 6,451 | 5.44% | 7,701 | 3.31% | 1.64 | 12,387 | 4.05% | 21,299 | 3.74% | 1.08 | 711 | 3.16% | 91 | 4.17% | 0.76 | 1.30 |
| Sleep Disorder | 3,786 | 3.19% | 6,198 | 2.66% | 1.20 | 5,684 | 1.86% | 11,843 | 2.08% | 0.89 | 405 | 1.80% | 57 | 2.61% | 0.69 | 1.02 |
| Depression | 7,259 | 6.13% | 11,733 | 5.04% | 1.22 | 11,085 | 3.62% | 23,114 | 4.06% | 0.89 | 728 | 3.23% | 111 | 5.09% | 0.63 | 1.02 |
| Anxiety | 9,686 | 8.17% | 16,256 | 6.98% | 1.17 | 18,538 | 6.06% | 39,293 | 6.91% | 0.88 | 895 | 3.97% | 110 | 5.04% | 0.79 | 1.00 |
| Joint Pain | 7,065 | 5.96% | 12,248 | 5.26% | 1.13 | 14,821 | 4.84% | 35,002 | 6.15% | 0.79 | 1,245 | 5.53% | 111 | 5.09% | 1.09 | 0.95 |
| Mood Changes | 521 | 0.44% | 1219 | 0.52% | 0.85 | 940 | 0.31% | 2,395 | 0.42% | 0.74 | 31 | 0.14% | 5 | 0.23% | 0.61 | 0.78 |
| Cough | 7,316 | 6.17% | 13,579 | 5.83% | 1.06 | 10,052 | 3.28% | 35,441 | 6.23% | 0.53 | 598 | 2.65% | 217 | 9.95% | 0.27 | 0.71 |
| Rash | 2,501 | 2.11% | 8030 | 3.45% | 0.61 | 7,348 | 2.40% | 27,922 | 4.91% | 0.49 | 560 | 2.49% | 108 | 4.95% | 0.50 | 0.54 |
| Fever | 2,072 | 1.75% | 12,167 | 5.23% | 0.33 | 3,855 | 1.26% | 31,305 | 5.50% | 0.23 | 429 | 1.90% | 248 | 11.37% | 0.17 | 0.27 |

Additionally, the proportions of patients who had post-acute diagnoses or symptoms of lung disorder, chest pain, depression, anxiety or joint pain in the COVID-19 cohort were greater than 2% in each database.

## Discussion

We have reported one of the largest studies about the incidence of PASC, and related symptoms or conditions, in a cohort of patients infected with COVID-19 using claims and EHR data. The incidences of some outcomes were greater for the COVID-19 cohort compared to the influenza cohort. Therefore, long term sequelae of COVID-19 may be different from influenza. For example, we expect that the long-term anosmia prevalence will likely be greater for COVID-19 patients.

Additionally, we observed symptoms and diagnoses in the COVID-19 cohort at non-negligible rates. Given the global prevalence of the COVID-19 pandemic, those conditions could have a major public health impact. Specifically, the global burden of respiratory illness, musculoskeletal disease, and psychiatric disorders may increase as a consequence of the COVID-19 pandemic. These findings suggest that PASC is likely composed as a heterogeneous constellation of continuing symptoms that do not resolve, rare but unusual symptoms, and prevalent serious symptoms.

Alternative approaches to characterizing PASC have used self-reported symptom data. Although those approaches are comprehensive, self-reported data may be different from patient assessments by healthcare providers. We expect that the differences in these kinds of data may help explain why the incidences of outcomes in our study are lower than incidences reported in patient self-assessment studies^9,10,13^.

A limitation of our analysis is that we did not validate our phenotypes, and therefore measurement error is possible; however, we used the CDC description for PASC in developing our phenotype^15^. Also, patient attrition to primary care sites out of network may have contributed to bias in the EHR database.

We have demonstrated the feasibility of characterizing the natural history of post-acute COVID-19 infections on both EHR and claims databases. The implications of our analysis may lead to public health interventions that can reduce the global burden of long-term sequelae from the COVID-19 pandemic.

## Conclusions

We have presented one of the largest characterizations of post-acute sequelae of SARS-CoV-2 (PASC) to date, examining data from multiple disparate populations. Electronic healthcare record data can be used to characterize PASC; additional replication of our analysis on other databases could strengthen our findings.

## Data Availability

Optum EHR and MarketScan CCAE are deidentified datasets available for licensure by Optum and IBM, respectively. The research done with data from Columbia University Irving Medical Center was approved by the IRB.

## References

[1] Wang D, Hu B, Hu C. et al. Clinical characteristics of 138 hospitalized patients with 2019 novel coronavirus–infected pneumonia in Wuhan, China. JAMA (2020) 323, 1061–1069.

[2] Goyal P, Choi JJ, Pinheiro LC, et al. Clinical characteristics of covid-19 in New York City. N Engl J Med (2020) 382(24):2372–2374.

[3] Guan W, Ni ZY, Liang WH, et al. Clinical characteristics of coronavirus disease 2019 in China. N Engl J Med (2020) 382(18):1708–1720.

[4] Zhou, F., Yu T., Du R. et al. Clinical course and risk factors for mortality of adult inpatients with COVID-19 in Wuhan, China: a retrospective cohort study. Lancet (2020) 395, 1054–1062.

[5] Burn E, You SC, Sena AG, et. al. Deep phenotyping of 34,128 adult patients hospitalised with COVID-19 in an international network study. Nat Commun. (2020) 6;11(1):5009.

[6] Argenziano MG, Bruce SL, Slater CL etal. Characterization and clinical course of 1000 patients with coronavirus disease 2019 in New York: retrospective case series. BMJ (2020);369:m1996.

[7] Dani M, Taraborrrelli P, Torocastro M etal. Autonomic dysfunction in ‘long COVID’: rationale, physiology and management strategies. Clinical Medicine (2021) Vol 21, No 1: e63–7.

[8] Shan MX, Tran Y, Vu KT etal. Postacute inpatient rehabilitation for COVID-19. BMJ Case Rep (2020);13:e237406.

[9] Carfi A, Bernabei R, Landi F etal. Persistent Symptoms in Patients After Acute COVID-19. JAMA (2020);324(6):603–605.

[10] Mandal S, Barnett J, Brill SE et al. ‘Long-COVID’: a cross-sectional study of persisting symptoms, biomarker and imaging abnormalities following hospitalisation for COVID-19. Thorax (2020);0:1–3.

[11] Logue JK, Franko NM, McCulloch DJ etal. Sequelae in Adults at 6 Months After COVID-19 Infection. JAMA Network Open. (2021);4(2):e210830.

[12] Bellan M, Soddu D, Balbo PE etal. Respiratory and Psychophysical Sequelae Among Patients With COVID-19 Four Months After Hospital Discharge. JAMA Network Open. (2021);4(1):e2036142.

[13] Sudre CH, Murray B, Varsavsky T etal. Attributes and predictors of long COVID. Nat Med (2021).

[14] https://covid19.nih.gov/sites/default/files/2021-02/PASC-ROA-OTA-Recovery-Cohort-Studies.pdf.

[15] https://www.cdc.gov/coronavirus/2019-ncov/long-term-effects.html.

