## Supplementary Appendix#1 for "Characterizing Post-Acute Sequelae of SARS-CoV-2 Infection across Claims and Electronic Health Record Databases"

Supplementary Appendix#1: Description of Databases

*OHDSI:*

Observational Health Data Sciences and Informatics (OHDSI) is an international, open-science collaborative of more than 220 healthcare organizations with a mission to improve health through the use of large-scale observational research. OHDSI maintains the Observational Medical Outcome Partnership (OMOP) Common Data Model (CDM), which standardizes how clinical data is represented, enabling standardized analysis methods within the OHDSI network. The OMOP CDM's schema represents structured data such as patient demographics, visits, conditions, procedures, lab results, vitals, and medications.

*Databases:*

<u>CUIMC 2020q4</u>

Columbia University Irving Medical Center (CUIMC)'s anonymized OMOP database comprises a mixture of inpatient and outpatient visits, spans a time period of 4 decades, 1980s-December 2020 (2020q4), and represents a population of 6 million patients. The data in the OMOP database were extracted from CUIMC and New York-Presbyterian Hospital's electronic health record (EHR) systems. The database currently holds information about the person (demographics), visits (inpatient, outpatient, and emergency), conditions (billing diagnoses and problem lists), drugs (outpatient prescriptions and inpatient orders and administrations), devices, measurements (laboratory tests and vital signs), and other observations (allergies).

<u>IBM Market Scan CCAE</u>

IBM MarketScan® Commercial Claims and Encounters Database (CCAE) represent data from individuals enrolled in United States employer-sponsored insurance health plans. The data includes adjudicated health insurance claims (e.g. inpatient, outpatient, and outpatient pharmacy) as well as enrollment data from large employers and health plans who provide private healthcare coverage to employees, their spouses, and dependents. Additionally, it captures laboratory tests for a subset of the covered lives. This administrative claims database includes a variety of fee-for-service, preferred provider organizations, and capitated health plans. The major data elements contained within this database are outpatient pharmacy dispensing claims (coded with National Drug Codes (NDC), inpatient and outpatient medical claims which provide procedure codes (coded in CPT-4, HCPCs, ICD-9-CM or ICD-10-PCS) and diagnosis codes (coded in ICD-9-CM or ICD-10-CM). The data also contain selected laboratory test results (those sent to a contracted thirds-party laboratory service provider) for a non-random sample of the population (coded with Logical Observation Identifiers Names and Codes (LOINC) codes).

<u>Optum EHR</u>

Optum© de-identified Electronic Health Record Dataset is derived from dozens of healthcare provider organizations in the United States (that include more than 700 hospitals and 7,000 Clinics treating more than 103 million patients) receiving care in the United States. The medical record data includes clinical information, inclusive of prescriptions as prescribed and administered, lab results, vital signs, body measurements, diagnoses, procedures, and information derived from clinical Notes using Natural Language Processing (NLP).
