## Supplementary Appendix#2 for "Characterizing Post-Acute Sequelae of SARS-CoV-2 Infection across Claims and Electronic Health Record Databases"

Supplementary Appendix#2: Concept Set Codes

Acute Kidney Injury

| Code | Name | Vocabulary |
|---|---|---|
| 14669001 | Acute renal failure syndrome | SNOMED |
| 35455006 | Acute tubular necrosis | SNOMED |
| 111407006 | Hemolytic uremic syndrome | SNOMED |
| 51292008 | Hepatorenal syndrome | SNOMED |
| 429224003 | Acute renal failure due to acute cortical necrosis | SNOMED |
| 61503006 | Acute nephritis | SNOMED |
| 197769007 | Acute pyelonephritis with medullary necrosis | SNOMED |
| 236392004 | Rapidly progressive glomerulonephritis | SNOMED |
| 197689003 | Rapidly progressive nephritic syndrome | SNOMED |
| 200118004 | Post-delivery acute renal failure with postnatal problem | SNOMED |
| 197697005 | Rapidly progressive nephritic syndrome, diffuse crescentic glomerulonephritis | SNOMED |
| 733839001 | Postpartum acute renal failure | SNOMED |
| 197685009 | Acute nephritic syndrome, diffuse endocapillary proliferative glomerulonephritis | SNOMED |
| 197684008 | Acute nephritic syndrome, diffuse mesangial proliferative glomerulonephritis | SNOMED |
| 716864001 | Hemorrhagic fever with renal syndrome | SNOMED |
| 197683002 | Acute nephritic syndrome, diffuse membranous glomerulonephritis | SNOMED |
| 197686005 | Acute nephritic syndrome, diffuse mesangiocapillary glomerulonephritis | SNOMED |

| | | |
|---|---|---|
| 197682007 | Acute nephritic syndrome, focal and segmental glomerular lesions | SNOMED |
| 197693009 | Rapidly progressive nephritic syndrome, diffuse mesangial proliferative glomerulonephritis | SNOMED |
| 197696001 | Rapidly progressive nephritic syndrome, dense deposit disease | SNOMED |
| 197681000 | Acute nephritic syndrome, minor glomerular abnormality | SNOMED |
| 197691006 | Rapidly progressive nephritic syndrome, focal and segmental glomerular lesions | SNOMED |
| 422593004 | Acute renal failure due to ACE inhibitor | SNOMED |
| 31005002 | Hepatorenal syndrome due to a procedure | SNOMED |
| 197692004 | Rapidly progressive nephritic syndrome, diffuse membranous glomerulonephritis | SNOMED |
| 197585004 | Acute diffuse nephritis | SNOMED |
| 236429004 | Acute drug-induced renal failure | SNOMED |
| 32093003 | Acute drug-induced tubulointerstitial nephritis | SNOMED |
| 197584000 | Acute focal nephritis | SNOMED |
| 1.44E+16 | Acute injury of kidney | SNOMED |
| 733137002 | Acute kidney failure stage 1 | SNOMED |
| 733138007 | Acute kidney failure stage 2 | SNOMED |
| 733139004 | Acute kidney failure stage 3 | SNOMED |
| 1.05E+15 | Acute kidney injury due to acute tubular necrosis due to circulatory failure | SNOMED |
| 1.05E+15 | Acute kidney injury due to acute tubular necrosis due to circulatory failure with histological evidence | SNOMED |

| | | |
|---|---|---|
| 1.05E+15 | Acute kidney injury due to acute tubular necrosis due to hypovolaemia | SNOMED |
| 1.05E+15 | Acute kidney injury due to acute tubular necrosis due to hypovolaemia with histological evidence | SNOMED |
| 1.05E+15 | Acute kidney injury due to acute tubular necrosis due to sepsis | SNOMED |
| 1.05E+15 | Acute kidney injury due to acute tubular necrosis due to sepsis with histological evidence | SNOMED |
| 1.05E+15 | Acute kidney injury due to acute tubular necrosis with histological evidence | SNOMED |
| 722095005 | Acute kidney injury due to circulatory failure | SNOMED |
| 722096006 | Acute kidney injury due to hypovolemia | SNOMED |
| 722278006 | Acute kidney injury due to sepsis | SNOMED |
| 726541005 | Acute kidney injury due to trauma | SNOMED |
| 8.52E+14 | Acute kidney injury stage 1 | SNOMED |
| 8.52E+14 | Acute kidney injury stage 2 | SNOMED |
| 8.52E+14 | Acute kidney injury stage 3 | SNOMED |
| 88380005 | Acute milk alkali syndrome | SNOMED |
| 197687001 | Acute nephritic syndrome, dense deposit disease | SNOMED |
| 4.04E+13 | Acute nephritis due to another disorder | SNOMED |
| 58574008 | Acute nephropathy | SNOMED |
| 1.08E+15 | Acute nephrotic syndrome | SNOMED |
| 1.40E+14 | Acute nontraumatic kidney injury | SNOMED |
| 2.70E+16 | Acute on chronic kidney injury | Nebraska Lexicon |
| 702718005 | Acute on chronic tubulointerstitial nephritis | SNOMED |
| 194909006 | Acute pericarditis secondary to uremia | SNOMED |

| | | |
|---|---|---|
| 438783006 | Acute postoperative renal failure | SNOMED |
| 32801008 | Acute pyelitis | SNOMED |
| 14343001 | Acute pyelitis with renal medullary necrosis | SNOMED |
| 3999002 | Acute pyelitis without renal medullary necrosis | SNOMED |
| 197770008 | Acute pyonephrosis | SNOMED |
| 7448003 | Acute pyonephrosis with renal medullary necrosis | SNOMED |
| 85495007 | Acute pyonephrosis without renal medullary necrosis | SNOMED |
| 444794000 | Acute renal cortical necrosis | SNOMED |
| 3.69E+14 | Acute renal failure caused by contrast agent | SNOMED |
| 269257004 | Acute renal failure due to crush syndrome | SNOMED |
| 423533009 | Acute renal failure due to ischemia | SNOMED |
| 8.42E+14 | Acute renal failure due to non-traumatic rhabdomyolysis | SNOMED |
| 429489008 | Acute renal failure due to obstruction | SNOMED |
| 36225005 | Acute renal failure due to procedure | SNOMED |
| 8.48E+14 | Acute renal failure due to traumatic rhabdomyolysis | SNOMED |
| 1.46E+14 | Acute renal failure due to tubular necrosis | SNOMED |
| 71909003 | Acute renal failure following molar AND/OR ectopic pregnancy | SNOMED |
| 8.40E+14 | Acute renal failure induced by aminoglycoside | SNOMED |
| 8.43E+14 | Acute renal failure induced by animal toxin | SNOMED |
| 8.40E+14 | Acute renal failure induced by cisplatin | SNOMED |
| 8.40E+14 | Acute renal failure induced by cyclosporin A | SNOMED |
| 8.43E+14 | Acute renal failure induced by heavy metal | SNOMED |

| | | |
|---|---|---|
| 8.40E+14 | Acute renal failure induced by non-steroidal anti-inflammatory drug | SNOMED |
| 8.43E+14 | Acute renal failure induced by plant toxin | SNOMED |
| 8.45E+14 | Acute renal failure induced by poison | SNOMED |
| 8.45E+14 | Acute renal failure induced by radiographic contrast media | SNOMED |
| 8.45E+14 | Acute renal failure induced by solvent | SNOMED |
| 1.30E+14 | Acute renal failure on dialysis | SNOMED |
| 1.89E+16 | Acute renal failure secondary to lithium toxicity | Nebraska Lexicon |
| 430535006 | Acute renal failure with oliguria | SNOMED |
| 236424009 | Acute renal impairment | SNOMED |
| 723189000 | Acute renal insufficiency | SNOMED |
| 236503001 | Acute scleroderma renal crisis | SNOMED |
| 28637003 | Acute tubulointerstitial nephritis | SNOMED |
| 236496000 | Acute urate nephropathy | SNOMED |
| 236433006 | Acute-on-chronic renal failure | SNOMED |
| 789660001 | Atypical hemolytic uremic syndrome | SNOMED |
| 444976001 | Congenital hemolytic uremic syndrome | SNOMED |
| 23697004 | Crush syndrome | SNOMED |
| 373421000 | Diarrhea-associated hemolytic uremic syndrome | SNOMED |
| 373422007 | Diarrhea-negative hemolytic uremic syndrome | SNOMED |
| 17380002 | Failed attempted termination of pregnancy with acute renal failure | SNOMED |
| 62216007 | Familial arthrogryposis-cholestatic hepatorenal syndrome | SNOMED |
| 722721004 | Familial hemolytic uremic syndrome | SNOMED |

| | | |
|---|---|---|
| 8199003 | Focal embolic nephritis syndrome | SNOMED |
| 36568005 | Hemolytic uremic syndrome of childhood | SNOMED |
| 78209002 | Hemolytic uremic syndrome, adult type | SNOMED |
| 213231008 | Hepatorenal syndrome as a complication of care | SNOMED |
| 22846003 | Hepatorenal syndrome following delivery | SNOMED |
| 47330001 | Idiopathic acute tubulointerstitial nephritis | SNOMED |
| 445258009 | Idiopathic rapidly progressive glomerulonephritis | SNOMED |
| 33561005 | Illegal termination of pregnancy with acute renal failure | SNOMED |
| 4451004 | Illegal termination of pregnancy with renal tubular necrosis | SNOMED |
| 609455009 | Induced termination of pregnancy complicated by acute renal failure | SNOMED |
| 609472002 | Induced termination of pregnancy complicated by acute renal failure with oliguria | SNOMED |
| 609482001 | Induced termination of pregnancy complicated by renal tubular necrosis | SNOMED |
| 52342006 | Legal termination of pregnancy with acute renal failure | SNOMED |
| 20483002 | Legal termination of pregnancy with renal tubular necrosis | SNOMED |
| 236428007 | Nephrotoxic acute renal failure | SNOMED |
| 301814009 | Post-renal renal failure | SNOMED |
| 236427002 | Post-traumatic acute tubular necrosis | SNOMED |
| 236426006 | Postoperative acute tubular necrosis | SNOMED |

| | | |
|---|---|---|
| 239932005 | Primary pauci-immune necrotizing and crescentic glomerulonephritis | SNOMED |
| 236432001 | Pulmonary renal syndrome | SNOMED |
| 3.69E+14 | Rapidly progressive nephritic syndrome co-occurrent and due to membranoproliferative glomerulonephritis type III | SNOMED |
| 197694003 | Rapidly progressive nephritic syndrome, diffuse endocapillary proliferative glomerulonephritis | SNOMED |
| 197695002 | Rapidly progressive nephritic syndrome, diffuse mesangiocapillary glomerulonephritis | SNOMED |
| 197690007 | Rapidly progressive nephritic syndrome, minor glomerular abnormality | SNOMED |
| 236451007 | Renal ocular syndrome | SNOMED |
| 307309005 | Transient acute renal failure | SNOMED |
| 276586004 | Transient neonatal renal tubular acidosis | SNOMED |
| 2.35E+16 | Transient renal failure | Nebraska Lexicon |
| 236431008 | Traumatic anuria - crush syndrome | SNOMED |
| 418839003 | Tubulointerstitial nephritis with uveitis syndrome | SNOMED |

Adult Rash

| Code | Name | Vocabulary |
|---|---|---|
| 271807003 | Eruption | SNOMED |
| 11381005 | Acne | SNOMED |
| 64144002 | Pruritic rash | SNOMED |
| 33339001 | Psoriasis with arthropathy | SNOMED |
| 56454009 | Pityriasis versicolor | SNOMED |
| 49882001 | Viral exanthem | SNOMED |
| 4776004 | Lichen planus | SNOMED |
| 77252004 | Pityriasis rosea | SNOMED |
| 200892002 | Generalized skin eruption caused by drug and medicament | SNOMED |

| | | |
|---|---|---|
| 30242009 | Scarlet fever | SNOMED |
| 64540004 | Lichen planopilaris | SNOMED |
| 186668002 | Gammaherpesviral mononucleosis | SNOMED |
| 54792008 | Erythematosquamous dermatosis | SNOMED |
| 88233000 | Parapsoriasis | SNOMED |
| 186535001 | Eczema herpeticum | SNOMED |
| 111873003 | Measles without complication | SNOMED |
| 238746008 | Acne keloid | SNOMED |
| 73442001 | Stevens-Johnson syndrome | SNOMED |
| 240302002 | Erythroderma neonatorum | SNOMED |
| 403194002 | Solar erythema | SNOMED |
| 28926001 | Eruption due to drug | SNOMED |
| 34730008 | Primate erythroparvovirus 1 infection | SNOMED |
| 34630004 | Pityriasis | SNOMED |
| 200893007 | Localized skin eruption caused by drug and medicament | SNOMED |
| 402296004 | Pityriasis alba | SNOMED |
| 37042000 | Guttate psoriasis | SNOMED |
| 68266006 | Hypertrophic lichen planus | SNOMED |
| 54385001 | Exanthema subitum | SNOMED |
| 49706007 | Neonatal acne | SNOMED |
| 20343006 | Pigmented purpuric lichenoid dermatitis of Gougerot and Blum | SNOMED |
| 41890004 | Lichen nitidus | SNOMED |
| 58767000 | Toxic erythema | SNOMED |
| 42228007 | Acne conglobata | SNOMED |
| 23894009 | Acne varioliformis | SNOMED |
| 42442001 | Lichen striatus | SNOMED |
| 86487001 | Acute lichenoid pityriasis | SNOMED |
| 3755001 | Pityriasis rubra pilaris | SNOMED |
| 27520001 | Pustular psoriasis of palms and soles | SNOMED |
| 88697005 | Pruritic urticarial papules and plaques of pregnancy | SNOMED |
| 399992009 | Erythroderma | SNOMED |
| 23854007 | Small plaque parapsoriasis | SNOMED |
| 10065003 | Excoriated acne | SNOMED |

| | | |
|---|---|---|
| 768962006 | Lyell syndrome | SNOMED |
| 1.06E+16 | Psoriatic arthritis mutilans | SNOMED |
| 238612002 | Generalized pustular psoriasis | SNOMED |
| 73692007 | Fixed drug eruption | SNOMED |
| 402902002 | Roseola infantum | SNOMED |
| 19431000 | Rubella arthritis | SNOMED |
| 58872001 | Erythema elevatum diutinum | SNOMED |
| 65038009 | Fox-Fordyce disease | SNOMED |
| 38921001 | Measles with complication | SNOMED |
| 195900001 | Measles pneumonia | SNOMED |
| 109254000 | Lichenoid drug eruption | SNOMED |
| 1.25E+14 | Stevens Johnson syndrome AND toxic epidermal necrolysis overlap | SNOMED |
| 274119009 | Rubella in pregnancy | SNOMED |
| 10954006 | Tropical acne | SNOMED |
| 371627004 | ACE inhibitor-aggravated angioedema | SNOMED |
| 238995004 | Acneiform drug eruption | SNOMED |
| 199192005 | Maternal rubella during pregnancy - baby delivered | SNOMED |
| 403608009 | Serum sickness due to drug | SNOMED |
| 201002003 | Subacute active lichen planus | SNOMED |
| 16341002 | Parapsoriasis lichenoides | SNOMED |
| 89322006 | Urticaria medicamentosa | SNOMED |
| 68166002 | Keratitis in exanthema | SNOMED |
| 69047008 | Drug-induced photosensitivity | SNOMED |
| 55846006 | Xeroderma of eyelid | SNOMED |
| 186567003 | Rubella with neurological complication | SNOMED |
| 67081008 | Large plaque parapsoriasis | SNOMED |
| 83627000 | Phototoxic drug eruption | SNOMED |
| 239812005 | Psoriatic arthritis with distal interphalangeal joint involvement | SNOMED |
| 33090006 | Yabapox | SNOMED |
| 238752009 | Acne agminata | SNOMED |
| 72936003 | Acne atrophica | SNOMED |
| 17332000 | Acne cosmetica | SNOMED |
| 77816008 | Acne detergicans | SNOMED |
| 83218005 | Acne estivalis | SNOMED |

| | | |
|---|---|---|
| 4659007 | Acne fulminans | SNOMED |
| 403350000 | Acne fulminans with erythema nodosum | SNOMED |
| 87144001 | Acne indurata | SNOMED |
| 13101006 | Acne mechanica | SNOMED |
| 403359004 | Acne nodule | SNOMED |
| 22920003 | Acne of external chemical origin | SNOMED |
| 402641003 | Acne of external origin | SNOMED |
| 75867005 | Acne rosacea, papular type | SNOMED |
| 201224007 | Acne urticata | SNOMED |
| 42200001 | Acne venenata | SNOMED |
| 402642005 | Acne with gram negative folliculitis | SNOMED |
| 402644006 | Acneiform eruption | SNOMED |
| 402643000 | Acneiform eruption due to chemical | SNOMED |
| 238640007 | Acrokeratosis paraneoplastica of Bazex | SNOMED |
| 400018004 | Acrokeratosis verruciformis of Darier disease | SNOMED |
| 400085009 | Acrokeratosis verruciformis of Hopf | SNOMED |
| 239098009 | Acropustulosis of infancy | SNOMED |
| 200999007 | Actinic lichen planus | SNOMED |
| 402311006 | Actively extending plaque psoriasis | SNOMED |
| 721542002 | Acute blistering eruption of skin | SNOMED |
| 238550003 | Acute cheiropodopompholyx | SNOMED |
| 721543007 | Acute desquamating eruption of skin | SNOMED |
| 723015000 | Acute discoid eruption of skin | SNOMED |
| 723010005 | Acute eruption of skin | SNOMED |
| 723003004 | Acute eruptive lichen planus | SNOMED |
| 723014001 | Acute exudative skin eruption | SNOMED |
| 702617007 | Acute generalized exanthematous pustulosis | SNOMED |
| 402327004 | Acute generalized pustular flare of preexisting plaque psoriasis | SNOMED |

| | | |
|---|---|---|
| 402326008 | Acute generalized pustular psoriasis de novo | SNOMED |
| 402312004 | Acute guttate psoriasis | SNOMED |
| 723012002 | Acute maculopapular eruption of skin | SNOMED |
| 723013007 | Acute papular eruption of skin | SNOMED |
| 723011009 | Acute purpuric eruption of skin | SNOMED |
| 59663001 | Adult premenstrual acne | SNOMED |
| 40175007 | Allergic dermatitis due to bite of Ctenocephalides felis | SNOMED |
| 240854000 | Ancylostomal cutaneous larva migrans | SNOMED |
| 720493003 | Annular atrophic lichen planus | SNOMED |
| 402745002 | Annular erythema due to drug | SNOMED |
| 201000006 | Annular lichen planus | SNOMED |
| 707247006 | Annular oral lichen planus | SNOMED |
| 19514005 | Arthritis mutilans | SNOMED |
| 241960008 | Aspirin-induced angioedema-urticaria | SNOMED |
| 25858008 | Atrophic lichen planus | SNOMED |
| 238663002 | Atrophic oral lichen planus | SNOMED |
| 238619006 | Atypical adult pityriasis rubra pilaris | SNOMED |
| 238622008 | Atypical juvenile pityriasis rubra pilaris | SNOMED |
| 725148000 | Atypical lichen myxedematosus | SNOMED |
| 240483006 | Atypical measles | SNOMED |
| 403622009 | B-cell pseudolymphoma due to drug | SNOMED |
| 402858006 | Bowel-bypass syndrome | SNOMED |
| 53354003 | Bromide acne | SNOMED |
| 201215003 | Bromine acne | SNOMED |
| 8007005 | Bromoderma | SNOMED |
| 35189002 | Bullae AND sweat gland necrosis in drug-induced coma | SNOMED |
| 6111009 | Bullous lichen planus | SNOMED |
| 298139006 | Bullous weal | SNOMED |

| | | |
|---|---|---|
| 240656001 | Chagas' exanthem | SNOMED |
| 2.40E+14 | Chickenpox exanthem | SNOMED |
| 402328009 | Childhood pustular psoriasis | SNOMED |
| 83684005 | Chlorine acne | SNOMED |
| 31922003 | Chronic generalized exfoliative dermatitis | SNOMED |
| 402309002 | Chronic guttate pattern psoriasis | SNOMED |
| 402307000 | Chronic large plaque psoriasis | SNOMED |
| 402349005 | Chronic lichen planus | SNOMED |
| 711435001 | Chronic migratory panniculitis | SNOMED |
| 238587006 | Chronic prurigo | SNOMED |
| 402308005 | Chronic small plaque psoriasis | SNOMED |
| 402310007 | Chronic stable plaque psoriasis | SNOMED |
| 238617008 | Circinate and annular pustular psoriasis | SNOMED |
| 238621001 | Circumscribed juvenile pityriasis rubra pilaris | SNOMED |
| 238618003 | Classical adult pityriasis rubra pilaris | SNOMED |
| 238620000 | Classical juvenile pityriasis rubra pilaris | SNOMED |
| 247468003 | Closed comedone | SNOMED |
| 201218001 | Colloid acne | SNOMED |
| 238744006 | Comedonal acne | SNOMED |
| 247467008 | Comedone | SNOMED |
| 238645002 | Confluent lichen planus | SNOMED |
| 722391005 | Congenital lethal erythroderma | SNOMED |
| 59721007 | Congenital syphilitic pemphigus | SNOMED |
| 240561005 | Corona veneris | SNOMED |
| 240546009 | Coxsackie virus exanthem | SNOMED |
| 19362000 | Cutaneous larva migrans | SNOMED |
| 16429005 | Cutaneous larva migrans by Ancylostoma braziliense | SNOMED |
| 73532000 | Cutaneous larva migrans by Ancylostoma caninum | SNOMED |

| | | |
|---|---|---|
| 442081006 | Cutaneous larva migrans due to Uncinaria | SNOMED |
| 240855004 | Cutaneous strongyloidiasis | SNOMED |
| 13277001 | Cystic acne | SNOMED |
| 784339002 | Deficiency of interleukin 36 receptor antagonist | SNOMED |
| 371062005 | Demodex acne | SNOMED |
| 238588001 | Dermographic prurigo | SNOMED |
| 2.80E+16 | Diffuse papular eruption | Nebraska Lexicon |
| 402298003 | Diffuse pityriasis alba | SNOMED |
| 81856009 | Disseminate infundibulo-folliculitis | SNOMED |
| 238813009 | Drug exanthem | SNOMED |
| 241958006 | Drug-aggravated angioedema-urticaria | SNOMED |
| 238992001 | Drug-induced erythroderma | SNOMED |
| 238997007 | Drug-induced ichthyosiform reaction | SNOMED |
| 403626007 | Drug-induced pellagra | SNOMED |
| 277805004 | Drug-induced persistent light reaction | SNOMED |
| 238994000 | Drug-induced pseudolymphomatous eruption | SNOMED |
| 403609001 | Drug-induced Stevens-Johnson syndrome | SNOMED |
| 297941009 | Drug-induced toxic erythema | SNOMED |
| 238603004 | Eczematized psoriasis | SNOMED |
| 240482001 | Enteroviral exanthem | SNOMED |
| 77470003 | Enteroviral exanthematous fever | SNOMED |
| 238667001 | Erosive lichen planus of lips | SNOMED |
| 721212001 | Erosive lichen planus of vagina | SNOMED |
| 238669003 | Erosive lichen planus of vulva | SNOMED |
| 238662007 | Erosive oral lichen planus | SNOMED |
| 238733003 | Erosive pustular dermatosis of the scalp | SNOMED |
| 713528004 | Eruption of skin caused by antiretroviral drug | SNOMED |
| 713316008 | Eruption of skin co-occurrent with human | SNOMED |

| | | |
|---|---|---|
| | immunodeficiency virus infection | |
| 403868009 | Eruptive basal cell papillomata | SNOMED |
| 238711000 | Eruptive lentiginosis | SNOMED |
| 77300003 | Erythema gyratum repens | SNOMED |
| 22972008 | Erythema multiforme, dermal type | SNOMED |
| 73658009 | Erythema multiforme, epidermal type | SNOMED |
| 86563000 | Erythema multiforme, mixed dermal-epidermal type | SNOMED |
| 76097009 | Erythema nodosum migrans | SNOMED |
| 763767006 | Erythema palmare hereditarium | SNOMED |
| 204919003 | Erythematous pinta plaque | SNOMED |
| 403620001 | Erythroderma due to vancomycin | SNOMED |
| 733206005 | Erythroderma in infancy | SNOMED |
| 402300003 | Erythroderma of unknown etiology | SNOMED |
| 78631004 | Erythromelanosis follicularis of face AND/OR neck | SNOMED |
| 735958007 | Exanthem caused by Chlamydophila psittaci | SNOMED |
| 423333008 | Exanthem due to chicken pox | SNOMED |
| 428633000 | Exanthem due to herpes zoster | SNOMED |
| 424306000 | Exanthem due to measles virus | SNOMED |
| 238812004 | Exanthematous disorder | SNOMED |
| 43591003 | Exanthematous infectious disease | SNOMED |
| 239099001 | Facial Afro-Caribbean childhood eruption | SNOMED |
| 403209006 | Familial actinic prurigo | SNOMED |
| 403210001 | Familial actinic prurigo of lip | SNOMED |
| 402860008 | Febrile ulceronecrotic pityriasis lichenoides acuta | SNOMED |
| 22583005 | Fixed drug eruption due to phenolphthalein | SNOMED |
| 400108007 | Flexural lichen planus | SNOMED |
| 238600001 | Flexural psoriasis | SNOMED |

| | | |
|---|---|---|
| 403372003 | Fox-Fordyce disease of axillae | SNOMED |
| 403373008 | Fox-Fordyce disease of vulva | SNOMED |
| 402292002 | Frictional lichenoid eruption | SNOMED |
| 717055000 | Frontal fibrosing alopecia | SNOMED |
| 123705009 | Generalized exfoliative contact dermatitis | SNOMED |
| 238613007 | Generalized pustular psoriasis of pregnancy | SNOMED |
| 38360005 | Generalized pustular psoriasis of von Zumbush | SNOMED |
| 28840001 | Generalized pustular psoriasis, exanthematous type | SNOMED |
| 725119006 | Generalized rash | SNOMED |
| 238668006 | Genital lichen planus | SNOMED |
| 240467006 | Genital Molluscum contagiosum | SNOMED |
| 233625007 | Giant cell pneumonia | SNOMED |
| 403357002 | Giant comedo | SNOMED |
| 238632001 | Giant porokeratosis | SNOMED |
| 240860000 | Gnathostomal cutaneous larva migrans | SNOMED |
| 402958005 | Gonococcal bacteremia-induced pustular vasculitis | SNOMED |
| 718215008 | Graham Little Piccardi Lassueur syndrome | SNOMED |
| 238682006 | Granuloma multiforme | SNOMED |
| 397845003 | Granulomatous rosacea | SNOMED |
| 22818000 | Granulosis rubra nasi | SNOMED |
| 402313009 | Guttate flare of psoriasis with preexisting plaques | SNOMED |
| 238647005 | Guttate lichen planus | SNOMED |
| 2176006 | Halogen acne | SNOMED |
| 403624005 | Halogen eruption | SNOMED |
| 70139002 | Harara | SNOMED |
| 201016008 | Hebra's prurigo | SNOMED |
| 240485004 | Hemorrhagic rubella | SNOMED |
| 2.49E+15 | Herpes zoster of skin of neck | SNOMED |
| 247466004 | Herpetiform eruption | SNOMED |
| 403354009 | Hydration acne | SNOMED |
| 200832000 | Hydroa estivale | SNOMED |
| 238664008 | Hypertrophic oral lichen planus | SNOMED |

| | | |
|---|---|---|
| 721171007 | Hypertrophic vulval lichen planus | SNOMED |
| 201219009 | Infantile acne | SNOMED |
| 238616004 | Infantile pustular psoriasis | SNOMED |
| 238815002 | Infectious mononucleosis ampicillin reaction | SNOMED |
| 238370005 | Infectious mononucleosis exanthem | SNOMED |
| 428101005 | Inflammatory acne | SNOMED |
| 32226008 | Iodide acne | SNOMED |
| 201217006 | Iodine acne | SNOMED |
| 89414003 | Iododerma | SNOMED |
| 410482007 | Iritis in psoriatic arthritis | SNOMED |
| 47277009 | Izumi fever | SNOMED |
| 238743000 | Juvenile acne | SNOMED |
| 239802003 | Juvenile psoriatic arthritis | SNOMED |
| 239803008 | Juvenile psoriatic arthritis with psoriasis | SNOMED |
| 239804002 | Juvenile psoriatic arthritis without psoriasis | SNOMED |
| 238615000 | Juvenile pustular psoriasis | SNOMED |
| 238609000 | K√É¬∂bner psoriasis | SNOMED |
| 402348002 | K√É¬∂bner reaction from lichen planus | SNOMED |
| 238628007 | Keratosis circumscripta | SNOMED |
| 238611009 | Lapiere type of psoriasis | SNOMED |
| 102606000 | Leser-Tr√É¬©lat sign | SNOMED |
| 721180007 | Lichen planopilaris of vulva | SNOMED |
| 726476005 | Lichen planus co-occurrent with onycholysis | SNOMED |
| 201001005 | Lichen planus obtusus | SNOMED |
| 718068005 | Lichen planus of gingiva | SNOMED |
| 238671003 | Lichen planus of glans penis | SNOMED |
| 238666005 | Lichen planus of lips | SNOMED |
| 238658001 | Lichen planus of nail | SNOMED |
| 238656002 | Lichen planus of palms and soles | SNOMED |
| 238670002 | Lichen planus of penis | SNOMED |
| 238655003 | Lichen planus of scalp | SNOMED |
| 238665009 | Lichen planus of tongue | SNOMED |
| 237112004 | Lichen planus of vulva | SNOMED |
| 238653005 | Lichen planus pemphigoides | SNOMED |
| 717061002 | Lichen planus pigmentosus | SNOMED |

| | | |
|---|---|---|
| 238652000 | Lichen planus-lupus erythematosus overlap | SNOMED |
| 3358001 | Lichen ruber moniliformis | SNOMED |
| 724465009 | Lichen simplex of male genitalia | SNOMED |
| 4859009 | Lichen spinulosus | SNOMED |
| 373603008 | Lichen verrucosus et reticularis | SNOMED |
| 238530002 | Light - exacerbated acne | SNOMED |
| 44509000 | Linear lichen planus | SNOMED |
| 51696001 | Lipogranulomatosis subcutanea of Rothmann and Makai | SNOMED |
| 248413004 | Liver palms | SNOMED |
| 724877007 | Localized eruption of skin | SNOMED |
| 402297008 | Localized pityriasis alba | SNOMED |
| 81271001 | Localized pustular psoriasis | SNOMED |
| 403358007 | Macrocomedone | SNOMED |
| 238453003 | Maculae ceruleae | SNOMED |
| 271756005 | Macular eruption | SNOMED |
| 240558009 | Macular syphilide | SNOMED |
| 238814003 | Maculopapular drug eruption | SNOMED |
| 247471006 | Maculopapular eruption | SNOMED |
| 240677003 | Maculopapular yaws | SNOMED |
| 199194006 | Maternal rubella during pregnancy - baby not yet delivered | SNOMED |
| 199193000 | Maternal rubella in the puerperium - baby delivered during current episode of care | SNOMED |
| 199195007 | Maternal rubella in the puerperium - baby delivered during previous episode of care | SNOMED |
| 2.40E+14 | Measles virus exanthem | SNOMED |
| 186562009 | Measles with intestinal complications | SNOMED |
| 238425003 | Meningococcal rash | SNOMED |
| 238646001 | Micropapular lichen planus | SNOMED |
| 298138003 | Micropapular weal | SNOMED |
| 240484000 | Modified measles | SNOMED |

| | | |
|---|---|---|
| 397515005 | Molluscum contagiosum blepharoconjunctivitis | SNOMED |
| 765782003 | Molluscum contagiosum infection of eyelid | SNOMED |
| 765783008 | Molluscum contagiosum skin infection | SNOMED |
| 247470007 | Morbilliform eruption | SNOMED |
| 301447009 | Multimorphic rash | SNOMED |
| 403356006 | Multiple "sandpaper" type comedones | SNOMED |
| 238660004 | Mutilating lichen planus of fingers and toes | SNOMED |
| 402340009 | Nail dystrophy due to pityriasis rubra pilaris | SNOMED |
| 238659009 | Nail pterygium in lichen planus | SNOMED |
| 762543009 | Necrolytic acral erythema | SNOMED |
| 15576007 | Necrolytic migratory erythema | SNOMED |
| 232242004 | Neurodermatitis of external ear | SNOMED |
| 111200005 | Nodular elastosis with cysts and comedones | SNOMED |
| 238745007 | Nodulocystic acne | SNOMED |
| 403610006 | Non-pigmenting fixed drug eruption | SNOMED |
| 724511006 | Nonerosive lichen planus of oral mucosa | SNOMED |
| 241959003 | NSAID-induced angioedema-urticaria | SNOMED |
| 275955005 | O/E - allergic rash | SNOMED |
| 164375004 | O/E - blackheads | SNOMED |
| 164358005 | O/E - deep seated pustules | SNOMED |
| 164427005 | O/E - discoid rash | SNOMED |
| 391168003 | O/E - dribble rash | SNOMED |
| 164359002 | O/E - follicular pustules | SNOMED |
| 304386008 | O/E - itchy rash | SNOMED |
| 164328004 | O/E - macules | SNOMED |
| 164330002 | O/E - macules present | SNOMED |
| 164357000 | O/E - purulent pustules | SNOMED |
| 164354007 | O/E - pustules | SNOMED |
| 164356009 | O/E - pustules present | SNOMED |
| 395122007 | O/E - scalp rash | SNOMED |

| | | |
|---|---|---|
| 164361006 | O/E - weals present | SNOMED |
| 201220003 | Occupational acne | SNOMED |
| 403351001 | Oil acne | SNOMED |
| 271763005 | Open comedone | SNOMED |
| 235049008 | Oral lichen planus | SNOMED |
| 238661000 | Oral Wickham's striae in lichen planus | SNOMED |
| 56940005 | Palmar erythema | SNOMED |
| 271757001 | Papular eruption | SNOMED |
| 239100009 | Papular eruption of blacks | SNOMED |
| 397741002 | Papular eruption of chin | SNOMED |
| 707248001 | Papular oral lichen planus | SNOMED |
| 240560006 | Papular syphilide | SNOMED |
| 200766001 | Parakeratosis | SNOMED |
| 200981004 | Parapsoriasis atrophicans | SNOMED |
| 56253000 | Parapsoriasis en plaques | SNOMED |
| 200982006 | Parapsoriasis herpetiformis | SNOMED |
| 200984007 | Parapsoriasis papulata | SNOMED |
| 200985008 | Parapsoriasis varioliformis | SNOMED |
| 8.17E+15 | Parvovirus B19 infection complicating pregnancy | Nebraska Lexicon |
| 402238001 | Pebbly lichenification | SNOMED |
| 277785006 | Penicillin-induced angioedema-urticaria | SNOMED |
| 2.48E+15 | Penile eruption | Nebraska Lexicon |
| 1.08E+16 | Perianal rash | Nebraska Lexicon |
| 240469009 | Perinatal varicella | SNOMED |
| 276719000 | Phototherapy skin rash | SNOMED |
| 238629004 | Phrynoderma | SNOMED |
| 110989007 | Pigmented peribuccal erythrosis of Brocq | SNOMED |
| 403352008 | Pitch acne | SNOMED |
| 238924007 | Pityriasis amiantacea | SNOMED |
| 400201008 | Pityriasis capitis | SNOMED |
| 200993008 | Pityriasis folliculorum | SNOMED |
| 238989000 | Pityriasis rosea-like drug eruption | SNOMED |
| 200948000 | Pityriasis rubra | SNOMED |
| 200767005 | Pityriasis simplex | SNOMED |
| 778075008 | Pityriasis simplex capitis | SNOMED |
| 403426003 | Pityriasis steatoides | SNOMED |
| 200994002 | Pityriasis streptogenes | SNOMED |

| | | |
|---|---|---|
| 200965009 | Plaque psoriasis | SNOMED |
| 403355005 | Polyporous comedone | SNOMED |
| 80432009 | Porokeratosis of Mibelli | SNOMED |
| 9031009 | Porokeratosis of Mibelli, linear unilateral type | SNOMED |
| 20110000 | Porokeratosis of Mibelli, plaque type | SNOMED |
| 30005005 | Porokeratosis of Mibelli, superficial disseminated type | SNOMED |
| 238649008 | Post-inflammatory hyperpigmentation in lichen planus | SNOMED |
| 67896006 | Post-kala-azar dermal leishmaniasis | SNOMED |
| 403325003 | Postmenopausal frontal fibrosing alopecia | SNOMED |
| 404102002 | Premycotic eruption | SNOMED |
| 238546005 | Pruriginous atopic dermatitis | SNOMED |
| 201017004 | Prurigo mitis | SNOMED |
| 239103006 | Prurigo of pregnancy | SNOMED |
| 43118004 | Prurigo papule | SNOMED |
| 238589009 | Prurigo pigmentosa | SNOMED |
| 55608001 | Prurigo simplex | SNOMED |
| 42570001 | Pruritus senilis | SNOMED |
| 402748000 | Pseudolymphomatous eruption due to drug | SNOMED |
| 52230004 | Psoriasiform dermatitis | SNOMED |
| 238988008 | Psoriasiform drug eruption | SNOMED |
| 200962007 | Psoriasis annularis | SNOMED |
| 200963002 | Psoriasis circinata | SNOMED |
| 200964008 | Psoriasis diffusa | SNOMED |
| 200966005 | Psoriasis geographica | SNOMED |
| 200967001 | Psoriasis gyrata | SNOMED |
| 200968006 | Psoriasis inveterata | SNOMED |
| 200972005 | Psoriasis punctata | SNOMED |
| 200974006 | Psoriasis universalis | SNOMED |
| 238602009 | Psoriasis-eczema overlap condition | SNOMED |
| 239813000 | Psoriatic dactylitis | SNOMED |
| 724854007 | Purpura of skin and or skin-associated mucous membrane co-occurrent and due to coagulation disorder | SNOMED |

| | | |
|---|---|---|
| 724855008 | Purpura of skin caused by mechanical force | SNOMED |
| 284078000 | Purpuric rash | SNOMED |
| 59172008 | Pustular acne | SNOMED |
| 3533007 | Pustular bacterid | SNOMED |
| 200973000 | Pustular psoriasis | SNOMED |
| 238614001 | Pustular psoriasis in children | SNOMED |
| 784328000 | Pustular psoriasis of palm of hand | SNOMED |
| 784327005 | Pustular psoriasis of sole of foot | SNOMED |
| 240562003 | Pustular syphilide | SNOMED |
| 402663007 | Pustular vasculitis | SNOMED |
| 271760008 | Pustule | SNOMED |
| 29909004 | Pyoderma faciale | SNOMED |
| 785724007 | Pyoderma gangrenosum, acne, suppurative hidradenitis syndrome | SNOMED |
| 724015007 | Pyogenic arthritis, pyoderma gangrenosum, acne syndrome | SNOMED |
| 827051009 | Rash due to dribbling from mouth | SNOMED |
| 827052002 | Rash due to dribbling of urine | SNOMED |
| 442279002 | Rash of genitalia | SNOMED |
| 1.47E+16 | Rash of groin | SNOMED |
| 827044000 | Rash of scalp | SNOMED |
| 266128007 | Rash of secondary syphilis | SNOMED |
| 95332009 | Rash of systemic lupus erythematosus | SNOMED |
| 238644003 | Recurrent focal palmar peeling | SNOMED |
| 707249009 | Reticular oral lichen planus | SNOMED |
| 402903007 | Roseola infantum | SNOMED |
| 128191000 | Rubella encephalomyelitis | SNOMED |
| 1.08E+16 | Rubella in mother complicating childbirth | SNOMED |
| 84939004 | Rubella in mother complicating pregnancy, childbirth AND/OR puerperium | SNOMED |
| 297958004 | Rubelliform eruption | SNOMED |
| 61099007 | Rubeola scarlatinosa | SNOMED |

| | | |
|---|---|---|
| 200969003 | Rupioid psoriasis | SNOMED |
| 60684003 | SAPHO syndrome | SNOMED |
| 419735006 | Scaling eczema | SNOMED |
| 238608008 | Scalp psoriasis | SNOMED |
| 25847004 | Seborrheic psoriasis | SNOMED |
| 402239009 | Secondary lichenification | SNOMED |
| 400051008 | Senile asteatotic eczema | SNOMED |
| 213323001 | Serum rash | SNOMED |
| 774211005 | Severe dermatitis, multiple allergies, metabolic wasting syndrome | SNOMED |
| 2.40E+14 | Shingles exanthem | SNOMED |
| 238654004 | Site-specific lichen planus | SNOMED |
| 420281004 | Skin rash associated with AIDS | SNOMED |
| 402177006 | Solar pruritus | SNOMED |
| 109251008 | Solar pruritus of elbows | SNOMED |
| 444100007 | Southern tick-associated rash illness | SNOMED |
| 27620007 | Spongiotic psoriasiform dermatitis | SNOMED |
| 201222006 | Steroid acne | SNOMED |
| 724833000 | Stevens-Johnson and toxic epidermal necrolysis overlap syndrome caused by drug | SNOMED |
| 768946000 | Stevens-Johnson syndrome, toxic epidermal necrolysis spectrum | SNOMED |
| 186357007 | Streptococcal sore throat with scarlatina | SNOMED |
| 737346001 | Streptococcal toxin-mediated perineal erythema | SNOMED |
| 240857007 | Strongyloidal cutaneous larva migrans | SNOMED |
| 240856003 | Strongyloidal ground itch | SNOMED |
| 6048002 | Subacute generalized exfoliative dermatitis | SNOMED |
| 238586002 | Subacute prurigo | SNOMED |
| 403347003 | Superficial acne vulgaris | SNOMED |
| 403348008 | Superficial inflammatory acne vulgaris | SNOMED |

| | | |
|---|---|---|
| 403349000 | Superficial mixed comedonal and inflammatory acne vulgaris | SNOMED |
| 402747005 | Sweet's disease due to drug | SNOMED |
| 238392009 | Sycosis cruris | SNOMED |
| 403623004 | T-cell pseudolymphoma due to drug | SNOMED |
| 402744003 | Toxic epidermal necrolysis due to drug | SNOMED |
| 402359006 | Toxic epidermal necrolysis due to graft-versus-host disease | SNOMED |
| 768961004 | Toxic epidermal necrolysis due to infection | SNOMED |
| 238816001 | Toxic urticated erythema | SNOMED |
| 403606008 | Toxic urticated erythema due to drug | SNOMED |
| 312106007 | Toxocara larva migrans of skin | SNOMED |
| 41156006 | Tuberculosis papulonecrotica | SNOMED |
| 238657006 | Ulcerative lichen planus of palms and soles | SNOMED |
| 51057008 | Ulerythema of cheeks | SNOMED |
| 400126005 | Ulerythema ophryogenes | SNOMED |
| 238601002 | Unstable psoriasis | SNOMED |
| 733110004 | Van den Bosch syndrome | SNOMED |
| 79893008 | Vesicular eruption | SNOMED |
| 277171005 | Vesicular eruption in ear canal | SNOMED |
| 95328003 | Vesicular skin eruptions of the temporal region | SNOMED |
| 28414003 | Vitamin A deficiency with xeroderma | SNOMED |
| 707250009 | Vulvovaginal gingival syndrome | SNOMED |
| 247472004 | Weal | SNOMED |
| 238651007 | Wickham's striae in lichen planus | SNOMED |
| 201025002 | Winter itch | SNOMED |
| 700298002 | Xeroderma of lower eyelid | SNOMED |
| 700346002 | Xeroderma of upper eyelid | SNOMED |
| 238648000 | Zosteriform lichen planus | SNOMED |

Alopecia

| Code | Name | Vocabulary |
|---|---|---|
| 238725004 | Non-scarring alopecia | SNOMED |
| 68225006 | Alopecia areata | SNOMED |
| 56317004 | Alopecia | SNOMED |
| 87872006 | Male pattern alopecia | SNOMED |
| 400088006 | Scarring alopecia | SNOMED |
| 86166000 | Alopecia universalis | SNOMED |
| 19754005 | Alopecia totalis | SNOMED |
| 27382006 | Alopecia mucinosa | SNOMED |
| 5860009 | Ophiasis | SNOMED |
| 2965006 | Congenital alopecia | SNOMED |
| 201146008 | Drug-induced androgenic alopecia | SNOMED |
| 238731001 | Pseudopelade | SNOMED |
| 59307008 | Syphilitic alopecia | SNOMED |
| 723005006 | Acute telogen effluvium | SNOMED |
| 720981000 | Alopecia and intellectual disability with hypergonadotropic hypogonadism syndrome | SNOMED |
| 783205005 | Alopecia antibody deficiency | SNOMED |
| 402637002 | Alopecia due to disturbance of hair cycle | SNOMED |
| 402639004 | Alopecia due to friction and trauma | SNOMED |
| 402638007 | Alopecia due to underlying disease | SNOMED |
| 201136006 | Alopecia febrilis | SNOMED |
| 201140002 | Alopecia follicularis | SNOMED |
| 201144006 | Alopecia hereditaria | SNOMED |
| 111024006 | Alopecia liminaris frontalis | SNOMED |
| 201137002 | Alopecia localis | SNOMED |
| 201141003 | Alopecia neurotica | SNOMED |
| 201134009 | Alopecia of pregnancy | SNOMED |
| 238729005 | Alopecia parvimaculata | SNOMED |
| 201143000 | Alopecia seborrheica | SNOMED |
| 201135005 | Alopecia senilis | SNOMED |
| 788417006 | Alopecia, epilepsy, intellectual disability syndrome Moynahan type | SNOMED |

| | | |
|---|---|---|
| 770941005 | Alopecia, progressive neurological defect, endocrinopathy syndrome | SNOMED |
| 720980004 | Alopecia, psychomotor epilepsy, periodontal pyorrhea, intellectual disability syndrome | SNOMED |
| 403318005 | Androgenetic alopecia in association with polycystic ovaries | SNOMED |
| 403798006 | Atrichia congenita | SNOMED |
| 715963002 | Atrichia with papular lesions | SNOMED |
| 719518004 | Autosomal dominant palmoplantar keratoderma and congenital alopecia | SNOMED |
| 719104003 | Autosomal recessive palmoplantar keratoderma and congenital alopecia syndrome | SNOMED |
| 403334008 | Brush alopecia | SNOMED |
| 403332007 | Brush roller alopecia | SNOMED |
| 201139004 | Cachectic alopecia | SNOMED |
| 1.09E+14 | Central centrifugal cicatricial alopecia | SNOMED |
| 720850008 | Choroidal atrophy and alopecia syndrome | SNOMED |
| 403335009 | Chronic diffuse alopecia | SNOMED |
| 403328001 | Chronic telogen effluvium | SNOMED |
| 403727008 | Cicatricial alopecia due to ionizing radiation | SNOMED |
| 402350005 | Cicatricial alopecia due to lichen planus | SNOMED |
| 403322000 | Circumscribed alopecia areata of beard area | SNOMED |
| 403321007 | Circumscribed alopecia areata of eyelashes/eyebrows | SNOMED |
| 403323005 | Circumscribed alopecia areata of limbs | SNOMED |
| 403320008 | Circumscribed alopecia areata of scalp | SNOMED |
| 403324004 | Circumscribed alopecia areata of trunk | SNOMED |

| | | |
|---|---|---|
| 77363004 | Concentric alopecia areata | SNOMED |
| 63935007 | Congenital absence of eyelash | SNOMED |
| 254225000 | Congenital alopecia with keratin cysts | SNOMED |
| 268288009 | Congenital generalized alopecia | SNOMED |
| 205592008 | Congenital localized alopecia | SNOMED |
| 201138007 | Diffuse alopecia | SNOMED |
| 46586006 | Diffuse alopecia areata | SNOMED |
| 73383004 | Drug-related alopecia | SNOMED |
| 771335004 | Ectodermal dysplasia syndactyly syndrome | SNOMED |
| 54539003 | Endocrine alopecia | SNOMED |
| 1108009 | Female pattern alopecia | SNOMED |
| 716088000 | Follicular hamartoma with alopecia and cystic fibrosis syndrome | SNOMED |
| 238469005 | Friction alopecia | SNOMED |
| 201132008 | Frontal alopecia of women | SNOMED |
| 717055000 | Frontal fibrosing alopecia | SNOMED |
| 725029001 | Frontonasal dysplasia with alopecia and genital anomaly syndrome | SNOMED |
| 31082002 | Frostbite alopecia | SNOMED |
| 722451006 | Gomez Lopez Hernandez syndrome | SNOMED |
| 721842008 | Hypogonadotropic hypogonadism with frontoparietal alopecia syndrome | SNOMED |
| 763404001 | Ichthyosis, alopecia, eclabion, ectropion, intellectual disability syndrome | SNOMED |
| 238728002 | Involutional alopecia | SNOMED |
| 238923001 | Lipedematous alopecia | SNOMED |
| 283872008 | Loss of outer third of eyebrow | SNOMED |
| 403331000 | Marginal alopecia | SNOMED |
| 403330004 | Massage alopecia | SNOMED |
| 65130004 | Nutritional alopecia | SNOMED |
| 275950000 | O/E - alopecia | SNOMED |
| 268921005 | O/E - patches of alopecia | SNOMED |

| | | |
|---|---|---|
| 763828007 | Odonto onycho dysplasia with alopecia syndrome | SNOMED |
| 239019002 | Odonto-onychial dysplasia with alopecia | SNOMED |
| 247541001 | Partial loss of hair | SNOMED |
| 716191002 | Perniola Krajewska Carnevale syndrome | SNOMED |
| 201142005 | Peroneal alopecia | SNOMED |
| 771186004 | Poikiloderma, alopecia, retrognathism, cleft palate syndrome | SNOMED |
| 59817009 | Postinfectional hypotrichosis | SNOMED |
| 403319002 | Postmenopausal androgenetic alopecia | SNOMED |
| 403325003 | Postmenopausal frontal fibrosing alopecia | SNOMED |
| 87038002 | Postpartum alopecia | SNOMED |
| 201133003 | Premature alopecia | SNOMED |
| 719275009 | Primary hypergonadotropic hypogonadism and partial alopecia syndrome | SNOMED |
| 733086003 | Pseudoprogeria syndrome | SNOMED |
| 22934003 | Radiation alopecia | SNOMED |
| 763630007 | Satoyoshi syndrome | SNOMED |
| 733204008 | Scarring alopecia due to traumatic injury | SNOMED |
| 722002002 | Scholte syndrome | SNOMED |
| 720345008 | Severe T-cell immunodeficiency, congenital alopecia, nail dystrophy syndrome | SNOMED |
| 403329009 | Short anagen syndrome | SNOMED |
| 721075001 | Short tarsus with absence of lower eyelashes syndrome | SNOMED |
| 403534007 | Sutural alopecia | SNOMED |
| 67772009 | Thermal burn alopecia | SNOMED |
| 771182002 | Thumb deformity, alopecia, pigmentation anomaly syndrome | SNOMED |
| 67488005 | Traumatic alopecia | SNOMED |
| 403535008 | Triangular alopecia | SNOMED |
| 238732008 | Tufted folliculitis | SNOMED |
| 403533001 | Vertical alopecia | SNOMED |

| | | |
|---|---|---|
| 816067005 | Woodhouse Sakati syndrome | SNOMED |

Altered Smell or Taste

| Code | Name | Domain |
|---|---|---|
| 2.61E+12 | Sensory disorder of smell and/or taste | Condition |
| 44169009 | Loss of sense of smell | Condition |
| 271801002 | Taste sense altered | Condition |
| 708673009 | Parosmia | Condition |
| 699656000 | Radiation induced taste impairment | Condition |
| 699220008 | Phantom taste | Condition |
| 722027009 | Kallman syndrome with heart disease | Condition |
| 771147003 | Isolated arhinencephaly | Condition |
| 37481009 | Anterior tongue taste disorder | Condition |
| 36905001 | Secondary bitter taste disorder | Condition |
| 66073004 | Glossopharyngeal taste disorder | Condition |
| 399993004 | Disorder of taste | Condition |
| 93559003 | Hypogonadism with anosmia | Condition |
| 67724008 | Secondary salt taste disorder | Condition |
| 348083004 | Primary taste disorder | Condition |
| 40307005 | Primary acid taste disorder | Condition |
| 41783004 | Traumatic anosmia | Condition |
| 47339000 | Secondary sweet taste disorder | Condition |
| 275462005 | Disorder of smell | Condition |
| 272028008 | C/O - anosmia | Condition |
| 34680005 | Primary sweet taste disorder | Condition |
| 247318005 | Abnormal taste in mouth | Condition |
| 164055003 | On examination - taste loss anterior 2/3 tongue | Condition |
| 20801005 | Secondary acid taste disorder | Condition |
| 15732002 | Secondary taste disorder | Condition |
| 230502003 | Congenital anosmia | Condition |
| 12869004 | Primary salt taste disorder | Condition |
| 112107000 | Primary bitter taste disorder | Condition |

Anxiety

| Code | Name | Vocabulary |
|---|---|---|
| 21897009 | Generalized anxiety disorder | SNOMED |
| 197480006 | Anxiety disorder | SNOMED |
| 47505003 | Posttraumatic stress disorder | SNOMED |
| 56576003 | Panic disorder without agoraphobia | SNOMED |
| 25501002 | Social phobia | SNOMED |
| 191736004 | Obsessive-compulsive disorder | SNOMED |
| 11806006 | Separation anxiety disorder of childhood | SNOMED |
| 35607004 | Panic disorder with agoraphobia | SNOMED |
| 67195008 | Acute stress disorder | SNOMED |
| 62351001 | Generalized social phobia | SNOMED |
| 17496003 | Organic anxiety disorder | SNOMED |
| 109006 | Anxiety disorder of childhood OR adolescence | SNOMED |
| 386810004 | Phobic disorder | SNOMED |
| 313182004 | Chronic post-traumatic stress disorder | SNOMED |
| 50026000 | Psychoactive substance-induced organic anxiety disorder | SNOMED |
| 82415003 | Agoraphobia without history of panic disorder without limited symptom attacks | SNOMED |
| 70691001 | Agoraphobia | SNOMED |
| 61569007 | Agoraphobia without history of panic disorder | SNOMED |
| 192042008 | Acute post-trauma stress state | SNOMED |
| 34938008 | Alcohol-induced anxiety disorder | SNOMED |
| 83253003 | Shyness disorder of childhood | SNOMED |
| 1686006 | Sedative, hypnotic AND/OR anxiolytic-induced anxiety disorder | SNOMED |
| 51493001 | Cocaine-induced anxiety disorder | SNOMED |
| 58535001 | Physical AND emotional exhaustion state | SNOMED |
| 192038005 | Acute fugue state due to acute stress reaction | SNOMED |
| 192037000 | Acute panic state due to acute stress reaction | SNOMED |
| 446175003 | Acute posttraumatic stress disorder following military combat | SNOMED |

| | | |
|---|---|---|
| 192041001 | Acute situational disturbance | SNOMED |
| 192039002 | Acute stupor state due to acute stress reaction | SNOMED |
| 191722009 | Agoraphobia with panic attacks | SNOMED |
| 1380006 | Agoraphobia without history of panic disorder with limited symptom attacks | SNOMED |
| 82339009 | Amphetamine-induced anxiety disorder | SNOMED |
| 724722007 | Anxiety disorder caused by dissociative drug | SNOMED |
| 2.26E+13 | Anxiety disorder caused by drug | SNOMED |
| 724723002 | Anxiety disorder caused by ketamine | SNOMED |
| 1.24E+16 | Anxiety disorder caused by methamphetamine | SNOMED |
| 724708007 | Anxiety disorder caused by methylenedioxymethamphetamine | SNOMED |
| 724654009 | Anxiety disorder caused by opioid | SNOMED |
| 762331007 | Anxiety disorder caused by stimulant | SNOMED |
| 737341006 | Anxiety disorder caused by synthetic cannabinoid | SNOMED |
| 762515000 | Anxiety disorder caused by synthetic cathinone | SNOMED |
| 52910006 | Anxiety disorder due to a general medical condition | SNOMED |
| 9.97E+15 | Anxiety disorder due to brain injury | Nebraska Lexicon |
| 1.07E+16 | Anxiety disorder in mother complicating childbirth | SNOMED |
| 37868008 | Anxiety disorder of adolescence | SNOMED |
| 53467004 | Anxiety disorder of childhood | SNOMED |
| 69479009 | Anxiety hyperventilation | SNOMED |
| 90790003 | Avoidant disorder of adolescence | SNOMED |
| 64165008 | Avoidant disorder of childhood | SNOMED |
| 37872007 | Avoidant disorder of childhood OR adolescence | SNOMED |
| 238976006 | Bromisodrophobia | SNOMED |
| 192611004 | Childhood phobic anxiety disorder | SNOMED |
| 699241002 | Chronic post-traumatic stress disorder following military combat | SNOMED |
| 426174008 | Chronic stress disorder | SNOMED |

| | | |
|---|---|---|
| 61157009 | Combat fatigue | SNOMED |
| 443919007 | Complex posttraumatic stress disorder | SNOMED |
| 191737008 | Compulsive neurosis | SNOMED |
| 446180007 | Delayed posttraumatic stress disorder following military combat | SNOMED |
| 111487009 | Dream anxiety disorder | SNOMED |
| 1.63E+16 | Mild major depressive disorder co-occurrent with anxiety single episode | SNOMED |
| 231504006 | Mixed anxiety and depressive disorder | SNOMED |
| 1.63E+16 | Moderate major depressive disorder co-occurrent with anxiety single episode | SNOMED |
| 428687006 | Nightmares associated with chronic post-traumatic stress disorder | SNOMED |
| 191738003 | Obsessional neurosis | SNOMED |
| 724693000 | Obsessive compulsive disorder caused by cocaine | SNOMED |
| 724730008 | Obsessive compulsive disorder caused by psychoactive substance | SNOMED |
| 762332000 | Obsessive compulsive disorder caused by stimulant | SNOMED |
| 762516004 | Obsessive compulsive disorder caused by synthetic cathinone | SNOMED |
| 10586006 | Occupation-related stress disorder | SNOMED |
| 723913009 | Olfactory reference disorder | SNOMED |
| 13438001 | Overanxious disorder of childhood | SNOMED |
| 371631005 | Panic disorder | SNOMED |
| 89948007 | Panic disorder with agoraphobia AND mild panic attacks | SNOMED |
| 8185002 | Panic disorder with agoraphobia AND moderate panic attacks | SNOMED |
| 59923000 | Panic disorder with agoraphobia AND panic attacks in full remission | SNOMED |
| 63909006 | Panic disorder with agoraphobia AND panic attacks in partial remission | SNOMED |
| 5509004 | Panic disorder with agoraphobia AND severe panic attacks | SNOMED |

| | | |
|---|---|---|
| 76868007 | Panic disorder with agoraphobia, agoraphobic avoidance in full remission AND mild panic attacks | SNOMED |
| 87798009 | Panic disorder with agoraphobia, agoraphobic avoidance in full remission AND moderate panic attacks | SNOMED |
| 11941006 | Panic disorder with agoraphobia, agoraphobic avoidance in full remission AND panic attacks in full remission | SNOMED |
| 111491004 | Panic disorder with agoraphobia, agoraphobic avoidance in full remission AND panic attacks in partial remission | SNOMED |
| 34116005 | Panic disorder with agoraphobia, agoraphobic avoidance in full remission AND severe panic attacks | SNOMED |
| 31781004 | Panic disorder with agoraphobia, agoraphobic avoidance in partial remission AND mild panic attacks | SNOMED |
| 32388005 | Panic disorder with agoraphobia, agoraphobic avoidance in partial remission AND moderate panic attacks | SNOMED |
| 22230001 | Panic disorder with agoraphobia, agoraphobic avoidance in partial remission AND panic attacks in full remission | SNOMED |
| 3158007 | Panic disorder with agoraphobia, agoraphobic avoidance in partial remission AND panic attacks in partial remission | SNOMED |
| 111490003 | Panic disorder with agoraphobia, agoraphobic avoidance in partial remission AND severe panic attacks | SNOMED |
| 63701002 | Panic disorder with agoraphobia, mild agoraphobic avoidance AND mild panic attacks | SNOMED |

| | | |
|---|---|---|
| 49564006 | Panic disorder with agoraphobia, mild agoraphobic avoidance AND moderate panic attacks | SNOMED |
| 24781009 | Panic disorder with agoraphobia, mild agoraphobic avoidance AND panic attacks in full remission | SNOMED |
| 50983008 | Panic disorder with agoraphobia, mild agoraphobic avoidance AND panic attacks in partial remission | SNOMED |
| 19766004 | Panic disorder with agoraphobia, mild agoraphobic avoidance AND severe panic attacks | SNOMED |
| 4932002 | Panic disorder with agoraphobia, moderate agoraphobic avoidance AND mild panic attacks | SNOMED |
| 82738004 | Panic disorder with agoraphobia, moderate agoraphobic avoidance AND moderate panic attacks | SNOMED |
| 64060000 | Panic disorder with agoraphobia, moderate agoraphobic avoidance AND panic attacks in full remission | SNOMED |
| 76812003 | Panic disorder with agoraphobia, moderate agoraphobic avoidance AND panic attacks in partial remission | SNOMED |
| 83631006 | Panic disorder with agoraphobia, moderate agoraphobic avoidance AND severe panic attacks | SNOMED |
| 1816003 | Panic disorder with agoraphobia, severe agoraphobic avoidance AND mild panic attacks | SNOMED |
| 30059008 | Panic disorder with agoraphobia, severe agoraphobic avoidance AND moderate panic attacks | SNOMED |
| 38328002 | Panic disorder with agoraphobia, severe agoraphobic avoidance AND panic attacks in full remission | SNOMED |
| 74010007 | Panic disorder with agoraphobia, severe agoraphobic avoidance AND panic attacks in partial remission | SNOMED |

| | | |
|---|---|---|
| 61212007 | Panic disorder with agoraphobia, severe agoraphobic avoidance AND severe panic attacks | SNOMED |
| 72861004 | Panic disorder without agoraphobia with mild panic attacks | SNOMED |
| 65064003 | Panic disorder without agoraphobia with moderate panic attacks | SNOMED |
| 82494000 | Panic disorder without agoraphobia with panic attacks in full remission | SNOMED |
| 53956006 | Panic disorder without agoraphobia with panic attacks in partial remission | SNOMED |
| 43150009 | Panic disorder without agoraphobia with severe panic attacks | SNOMED |
| 2.48E+14 | Paruresis | SNOMED |
| 55967005 | Phencyclidine-induced anxiety disorder | SNOMED |
| 403593004 | Phobic fear of skin cancer | SNOMED |
| 318784009 | Posttraumatic stress disorder, delayed onset | SNOMED |
| 192014006 | Psychogenic rumination | SNOMED |
| 1.63E+16 | Recurrent major depressive disorder co-occurrent with anxiety in full remission | SNOMED |
| 1.63E+16 | Recurrent major depressive disorder in partial remission co-occurrent with anxiety | SNOMED |
| 1.63E+16 | Recurrent mild major depressive disorder co-occurrent with anxiety | SNOMED |
| 1.63E+16 | Recurrent moderate major depressive disorder co-occurrent with anxiety | SNOMED |
| 1.63E+16 | Recurrent severe major depressive disorder co-occurrent with anxiety | SNOMED |
| 126943008 | Separation anxiety | SNOMED |
| 85061001 | Separation anxiety disorder of childhood, early onset | SNOMED |

| | | |
|---|---|---|
| 1.63E+16 | Severe major depressive disorder co-occurrent with anxiety single episode | SNOMED |
| 279611005 | Shell shock | SNOMED |
| 54587008 | Simple phobia | SNOMED |
| 2.12E+16 | Situational anxiety | Nebraska Lexicon |
| 191724005 | Social phobia, fear of eating in public | SNOMED |
| 191725006 | Social phobia, fear of public speaking | SNOMED |
| 191726007 | Social phobia, fear of public washing | SNOMED |
| 317816007 | Stockholm syndrome | SNOMED |
| 3.52E+14 | Stranger anxiety | SNOMED |
| 192044009 | Stress reaction causing mixed disturbance of emotion and conduct | SNOMED |
| 238966008 | Syphilophobia | SNOMED |
| 238965007 | Venereophobia | SNOMED |

Chest Pain

| Code | Name | Vocabulary |
|---|---|---|
| 29857009 | Chest pain | SNOMED |
| 194828000 | Angina pectoris | SNOMED |
| 4557003 | Preinfarction syndrome | SNOMED |
| 71884009 | Precordial pain | SNOMED |
| 16331000 | Heartburn | SNOMED |
| 1.60E+16 | Angina co-occurrent and due to coronary arteriosclerosis | SNOMED |
| 2237002 | Pleuritic pain | SNOMED |
| 1.60E+16 | Unstable angina co-occurrent and due to coronary arteriosclerosis | SNOMED |
| 274664007 | Chest pain on breathing | SNOMED |
| 239175003 | Post-thoracotomy pain syndrome | SNOMED |
| 87343002 | Prinzmetal angina | SNOMED |
| 59021001 | Angina decubitus | SNOMED |
| 735940001 | Pain of intercostal space | SNOMED |
| 83264000 | Epidemic pleurodynia | SNOMED |

| | | |
|---|---|---|
| 314116003 | Post infarct angina | SNOMED |
| 102587001 | Acute chest pain | SNOMED |
| 194823009 | Acute coronary insufficiency | SNOMED |
| 7.91E+11 | Angina associated with type 2 diabetes mellitus | SNOMED |
| 1.60E+16 | Angina co-occurrent and due to arteriosclerosis of coronary artery bypass graft | SNOMED |
| 61490001 | Angina, class I | SNOMED |
| 41334000 | Angina, class II | SNOMED |
| 85284003 | Angina, class III | SNOMED |
| 89323001 | Angina, class IV | SNOMED |
| 161973001 | Anterior chest wall pain | SNOMED |
| 7.92E+12 | Anterior pleuritic pain | SNOMED |
| 1.60E+16 | Arteriosclerosis of autologous arterial coronary artery bypass graft with angina | SNOMED |
| 1.60E+16 | Arteriosclerosis of autologous vein coronary artery bypass graft with angina | SNOMED |
| 371807002 | Atypical angina | SNOMED |
| 102589003 | Atypical chest pain | SNOMED |
| 1.57E+16 | Bilateral pain of shoulder blades | SNOMED |
| 426396005 | Cardiac chest pain | SNOMED |
| 233845001 | Cardiac syndrome X | SNOMED |
| 161972006 | Central chest pain | SNOMED |
| 279019008 | Central crushing chest pain | SNOMED |
| 9267009 | Chest pain at rest | SNOMED |
| 3.48E+13 | Chest pain due to pericarditis | SNOMED |
| 81953000 | Chest pain on exertion | SNOMED |
| 7.46E+15 | Chest pain rule out myocardial infarction | Nebraska Lexicon |
| 102588006 | Chest wall pain | SNOMED |
| 102591006 | Chest wall tenderness | SNOMED |
| 4.34E+14 | Chronic chest pain | SNOMED |
| 7.58E+15 | Costal chondral thoracic pain | Nebraska Lexicon |
| 161977000 | Costal margin chest pain | SNOMED |
| 59139008 | Crushing chest pain | SNOMED |
| 3368006 | Dull chest pain | SNOMED |
| 36859004 | Esophageal chest pain | SNOMED |

| | | |
|---|---|---|
| 300995000 | Exercise-induced angina | SNOMED |
| 722876002 | Functional heartburn | SNOMED |
| 95421005 | Intercostal myalgia | SNOMED |
| 247389006 | Intercostal neuralgia | SNOMED |
| 713839007 | Intercostal neuralgia as late effect of trauma | SNOMED |
| 230597003 | Intercostal post-herpetic neuralgia | SNOMED |
| 279048002 | Internal mammary artery syndrome | SNOMED |
| 225566008 | Ischemic chest pain | SNOMED |
| 285385002 | Left sided chest pain | SNOMED |
| 9.70E+13 | Localized chest pain | SNOMED |
| 281245003 | Musculoskeletal chest pain | SNOMED |
| 279036000 | Myofascial pain syndrome of thorax | SNOMED |
| 233821000 | New onset angina | SNOMED |
| 35928006 | Nocturnal angina | SNOMED |
| 274668005 | Non-cardiac chest pain | SNOMED |
| 1.57E+16 | Pain of bilateral sternoclavicular joints | SNOMED |
| 774135008 | Pain of left shoulder blade | SNOMED |
| 1.08E+15 | Pain of left sternoclavicular joint | SNOMED |
| 774134007 | Pain of right shoulder blade | SNOMED |
| 1.08E+15 | Pain of right sternoclavicular joint | SNOMED |
| 298731003 | Pain of sternum | SNOMED |
| 161974007 | Parasternal pain | SNOMED |
| 13057000 | Pleuropericardial chest pain | SNOMED |
| 713829001 | Postinfective intercostal neuralgia | SNOMED |
| 371806006 | Progressive angina | SNOMED |
| 10000006 | Radiating chest pain | SNOMED |
| 371810009 | Recurrent angina after coronary artery bypass graft | SNOMED |
| 371809004 | Recurrent angina after coronary stent placement | SNOMED |
| 371812001 | Recurrent angina after directional coronary atherectomy | SNOMED |

| | | |
|---|---|---|
| 371808007 | Recurrent angina after percutaneous transluminal coronary angioplasty | SNOMED |
| 371811008 | Recurrent angina post rotational atherectomy | SNOMED |
| 315025001 | Refractory angina | SNOMED |
| 4568003 | Retrosternal pain | SNOMED |
| 297217002 | Rib pain | SNOMED |
| 298740004 | Rib tender | SNOMED |
| 285386001 | Right sided chest pain | SNOMED |
| 20793008 | Scapulalgia | SNOMED |
| 5.38E+15 | Spasmodic midsternal pain | Nebraska Lexicon |
| 371030007 | Squeezing chest pain | SNOMED |
| 233819005 | Stable angina | SNOMED |
| 1.68E+16 | Stable angina due to coronary arteriosclerosis | SNOMED |
| 19057007 | Status anginosus | SNOMED |
| 202478007 | Sternoclavicular joint pain | SNOMED |
| 21470009 | Syncope anginosa | SNOMED |
| 103015000 | Thoracic nerve root pain | SNOMED |
| 429559004 | Typical angina | SNOMED |
| 1.60E+16 | Unstable angina co-occurrent and due to arteriosclerosis of coronary artery bypass graft | SNOMED |
| 1.60E+16 | Unstable angina due to arteriosclerosis of autologous vein coronary artery bypass graft | SNOMED |
| 285389008 | Upper chest pain | SNOMED |
| 89638008 | Xiphodynia | SNOMED |
| 89874002 | Xiphoidalgia syndrome | SNOMED |

Cough

| Code | Name | Vocabulary |
|---|---|---|
| 49727002 | Cough | SNOMED |
| 274708000 | Abnormal sputum | SNOMED |
| 8.11E+14 | [D]Episodic dry cough | SNOMED |
| 289113002 | Able to cough | SNOMED |
| 225576006 | Able to cough up sputum | SNOMED |
| 306780002 | Able to cough voluntarily | SNOMED |
| 275787002 | Acid-fast bacilli in sputum | SNOMED |
| 300959008 | Allergic cough | SNOMED |

| | | |
|---|---|---|
| 17986004 | Barking cough | SNOMED |
| 274399004 | Blood in sputum O/E | SNOMED |
| 6686005 | Blood streaked sputum | SNOMED |
| 61281005 | Bloodstained sputum | SNOMED |
| 62427007 | Bovine cough | SNOMED |
| 20670007 | Brassy cough | SNOMED |
| 2.05E+14 | Bronchial casts | SNOMED |
| 277910004 | Brown sputum | SNOMED |
| 272039006 | C/O - cough | SNOMED |
| 68154008 | Chronic cough | SNOMED |
| 248604008 | Clear sputum | SNOMED |
| 248589007 | Clearing throat - hawking | SNOMED |
| 365445003 | Color of sputum - finding | SNOMED |
| 365444004 | Consistency of sputum - finding | SNOMED |
| 248599002 | Copious sputum | SNOMED |
| 46789001 | Cough after eating | SNOMED |
| 7142008 | Cough at rest | SNOMED |
| 1.27E+16 | Cough due to ACE inhibitor | Nebraska Lexicon |
| 19282004 | Cough on exercise | SNOMED |
| 123819004 | Cough reflex absent | SNOMED |
| 102580004 | Cough suppression | SNOMED |
| 248593001 | Cough when swallowing | SNOMED |
| 135883003 | Cough with fever | SNOMED |
| 276314008 | Coughing ineffective | SNOMED |
| 277906002 | Creamy sputum | SNOMED |
| 289965001 | Croupy cough | SNOMED |
| 8.61E+14 | Dark green sputum | SNOMED |
| 52673003 | Decreased coughing | SNOMED |
| 289116005 | Difficulty coughing | SNOMED |
| 306782005 | Difficulty coughing voluntarily | SNOMED |
| 301245004 | Difficulty in coughing up sputum | SNOMED |
| 248605009 | Dirty sputum | SNOMED |
| 289114008 | Does cough | SNOMED |
| 301246003 | Does cough up sputum | SNOMED |
| 289115009 | Does not cough | SNOMED |
| 301247007 | Does not cough up sputum | SNOMED |
| 11833005 | Dry cough | SNOMED |
| 62618004 | Early morning cough | SNOMED |
| 301236000 | Effective cough | SNOMED |

| | | |
|---|---|---|
| 161933007 | Evening cough | SNOMED |
| 111987004 | Expectoration of currant jelly sputum | SNOMED |
| 301293005 | Finding of odor of sputum | SNOMED |
| 366124008 | Finding related to ability to cough | SNOMED |
| 366126005 | Finding related to ability to cough up sputum | SNOMED |
| 366125009 | Finding related to ability to cough voluntarily | SNOMED |
| 43516009 | Foul smelling sputum | SNOMED |
| 301290008 | Frank blood in sputum | SNOMED |
| 68423001 | Frothy sputum | SNOMED |
| 277900008 | Gray sputum | SNOMED |
| 277908001 | Green sputum | SNOMED |
| 59994004 | Hacking cough | SNOMED |
| 2.48E+16 | Increased sputum production | Nebraska Lexicon |
| 62731002 | Increasing frequency of cough | SNOMED |
| 248600004 | Moderate sputum | SNOMED |
| 161932002 | Morning cough | SNOMED |
| 64503007 | Mucoid sputum | SNOMED |
| 275718000 | Mucoid sputum - O/E | SNOMED |
| 8955008 | Mucopurulent sputum | SNOMED |
| 769211009 | No cough strength | SNOMED |
| 248602007 | No sputum | SNOMED |
| 62548007 | Nocturnal cough | SNOMED |
| 161947006 | Nocturnal cough / wheeze | SNOMED |
| 309651002 | O/E - sputum | SNOMED |
| 247410004 | Painful cough | SNOMED |
| 8.61E+14 | Pale green sputum | SNOMED |
| 43025008 | Paroxysmal cough | SNOMED |
| 284523002 | Persistent cough | SNOMED |
| 301289004 | Pluggy sputum | SNOMED |
| 111962006 | Postural cough | SNOMED |
| 445241004 | Postviral cough | SNOMED |
| 28743005 | Productive cough | SNOMED |
| 161923004 | Productive cough -clear sputum | SNOMED |
| 161924005 | Productive cough -green sputum | SNOMED |
| 161925006 | Productive cough-yellow sputum | SNOMED |

| | | |
|---|---|---|
| 301292000 | Profuse watery sputum | SNOMED |
| 42192008 | Purulent sputum | SNOMED |
| 274400006 | Pus in sputum O/E | SNOMED |
| 2.00E+14 | Reflux cough | SNOMED |
| 417850002 | Respiratory tract congestion and cough | SNOMED |
| 24816000 | Rusty sputum | SNOMED |
| 248601000 | Scanty sputum | SNOMED |
| 46802002 | Smokers' cough | SNOMED |
| 63000007 | Spasmodic cough | SNOMED |
| 167987009 | Sputum - not infected | SNOMED |
| 248596009 | Sputum - symptom | SNOMED |
| 271826006 | Sputum abnormal - amount | SNOMED |
| 271827002 | Sputum abnormal - color | SNOMED |
| 271828007 | Sputum abnormal - odor | SNOMED |
| 365443005 | Sputum appearance - finding | SNOMED |
| 167990003 | Sputum appears normal | SNOMED |
| 1.89E+16 | Sputum culture positive for extended beta lactam resistant E coli | Nebraska Lexicon |
| 168476005 | Sputum cytology | SNOMED |
| 168477001 | Sputum cytology positive | SNOMED |
| 275719008 | Sputum evidence of infection | SNOMED |
| 167986000 | Sputum examination: abnormal | SNOMED |
| 167985001 | Sputum examination: normal | SNOMED |
| 248595008 | Sputum finding | SNOMED |
| 758674005 | Sputum loose | SNOMED |
| 167996009 | Sputum microscopy: NAD | SNOMED |
| 277357006 | Sputum retention | SNOMED |
| 167999002 | Sputum: asbestos bodies | SNOMED |
| 168000008 | Sputum: blood cells present | SNOMED |
| 270031000 | Sputum: contains blood | SNOMED |
| 167997000 | Sputum: eosinophilia | SNOMED |
| 167991004 | Sputum: excessive - mucoid | SNOMED |
| 167998005 | Sputum: malignant cells | SNOMED |
| 168001007 | Sputum: pus cells present | SNOMED |
| 269912000 | Sputum: tubercle on Z-N stain | SNOMED |
| 277902000 | Stringy sputum | SNOMED |
| 301294004 | Sweet smelling sputum | SNOMED |
| 277903005 | Thick sputum | SNOMED |

| | | |
|---|---|---|
| 277904004 | Thin sputum | SNOMED |
| 248594007 | Tracheal esophageal fistula cough | SNOMED |
| 248590003 | Unable to cough | SNOMED |
| 225575005 | Unable to cough up sputum | SNOMED |
| 306781003 | Unable to cough voluntarily | SNOMED |
| 315246003 | Unexplained cough | SNOMED |
| 365446002 | Volume of sputum - finding | SNOMED |
| 301291007 | Watery sputum | SNOMED |
| 427931002 | White sputum | SNOMED |
| 277907006 | Yellow sputum | SNOMED |

COVID Conditions

| Code | Name | Vocabulary |
|---|---|---|
| 840539006 | Disease caused by 2019-nCoV | SNOMED |
| 840544004 | Suspected disease caused by 2019-nCoV | SNOMED |
| 186747009 | Coronavirus infection | SNOMED |
| 27619001 | Disease due to Coronaviridae | SNOMED |
| 398447004 | Severe acute respiratory syndrome | SNOMED |
| 441590008 | Pneumonia due to Severe acute respiratory syndrome coronavirus | SNOMED |
| 6.51E+11 | Middle East respiratory syndrome | SNOMED |
| 715882005 | Severe acute respiratory syndrome of upper respiratory tract | SNOMED |
| 1.24E+15 | Myocarditis caused by 2019 novel coronavirus | SNOMED |
| 1.24E+15 | Infection of upper respiratory tract caused by 2019 novel coronavirus | SNOMED |
| 1.24E+15 | Pneumonia caused by 2019 novel coronavirus | SNOMED |
| 1.24E+15 | Encephalopathy caused by 2019 novel coronavirus | SNOMED |
| 1.24E+15 | Gastroenteritis caused by 2019 novel coronavirus | SNOMED |
| 1.24E+15 | Otitis media caused by 2019 novel coronavirus | SNOMED |

| | | |
|---|---|---|
| 713084008 | Pneumonia caused by Human coronavirus | SNOMED |
| 408688009 | Healthcare associated severe acute respiratory syndrome | SNOMED |
| OMOP4873908 | Infection of lower respiratory tract due to COVID-19 | OMOP Extension |
| OMOP4873910 | Asymptomatic COVID-19 | OMOP Extension |
| OMOP4873911 | Acute respiratory distress syndrome (ARDS) due to COVID-19 | OMOP Extension |
| OMOP4873907 | Respiratory infection due to COVID-19 | OMOP Extension |
| OMOP4873909 | Bronchitis due to COVID-19 | OMOP Extension |
| OMOP4873906 | Acute bronchitis due to COVID-19 | OMOP Extension |

Dementia or Memory Impairment or Inattentiveness

| Code | Name | Vocabulary |
|---|---|---|
| 48167000 | Amnesia | SNOMED |
| 26929004 | Alzheimer's disease | SNOMED |
| 52448006 | Dementia | SNOMED |
| 191519005 | Dementia associated with another disease | SNOMED |
| 409083006 | Finding related to attentiveness | SNOMED |
| 191449005 | Uncomplicated senile dementia | SNOMED |
| 1.59E+12 | Dementia with behavioral disturbance | SNOMED |
| 40425004 | Postconcussion syndrome | SNOMED |
| 416975007 | Primary degenerative dementia of the Alzheimer type, senile onset | SNOMED |
| 1.63E+16 | Vascular dementia without behavioral disturbance | SNOMED |
| 70936005 | Multi-infarct dementia, uncomplicated | SNOMED |
| 191461002 | Senile dementia with delirium | SNOMED |
| 191451009 | Uncomplicated presenile dementia | SNOMED |
| 2.89E+14 | Vascular dementia with behavioral disturbance | SNOMED |

| | | |
|---|---|---|
| 230736007 | Transient global amnesia | SNOMED |
| 10349009 | Multi-infarct dementia with delirium | SNOMED |
| 416780008 | Primary degenerative dementia of the Alzheimer type, presenile onset | SNOMED |
| 371024007 | Senile dementia with delusion | SNOMED |
| 268612007 | Senile and presenile organic psychotic conditions | SNOMED |
| 191459006 | Senile dementia with depression | SNOMED |
| 191452002 | Presenile dementia with delirium | SNOMED |
| 191455000 | Presenile dementia with depression | SNOMED |
| 76039005 | Disturbance of attention | SNOMED |
| 14070001 | Multi-infarct dementia with depression | SNOMED |
| 22058002 | Inattention | SNOMED |
| 25772007 | Multi-infarct dementia with delusions | SNOMED |
| 3.11E+13 | Presenile dementia with delusions | SNOMED |
| 88822006 | Anterograde amnesia | SNOMED |
| 51928006 | General paresis - neurosyphilis | SNOMED |
| 12348006 | Presenile dementia | SNOMED |
| 429998004 | Vascular dementia | SNOMED |
| 51921000 | Retrograde amnesia | SNOMED |
| 288769005 | Able to direct attention | SNOMED |
| 716403002 | Able to divide attention | SNOMED |
| 716443007 | Able to shift attention | SNOMED |
| 716452003 | Able to sustain attention | SNOMED |
| 46991000 | Absent minded | SNOMED |
| 783258000 | ADan amyloidosis | SNOMED |
| 9.78E+13 | Altered behavior in Alzheimer's disease | SNOMED |
| 8.24E+13 | Altered behavior in Huntington's dementia | SNOMED |
| 1.42E+14 | Alzheimer's disease co-occurrent with delirium | SNOMED |
| 2.46E+14 | Alzheimers continuum | Nebraska Lexicon |

| | | |
|---|---|---|
| 247607004 | Amnesia for day to day facts | SNOMED |
| 247611005 | Amnesia for important personal information | SNOMED |
| 42176003 | Amnesia for recent events | SNOMED |
| 6149008 | Amnesia for remote events | SNOMED |
| 191464005 | Arteriosclerotic dementia with delirium | SNOMED |
| 191466007 | Arteriosclerotic dementia with depression | SNOMED |
| 191465006 | Arteriosclerotic dementia with paranoia | SNOMED |
| 247768004 | Attends to adults' choice of activity | SNOMED |
| 1.97E+14 | Attention delay | SNOMED |
| 1.62E+16 | Behavioral disturbance co-occurrent and due to late onset Alzheimer dementia | SNOMED |
| 1.42E+14 | Cognitive deficit in attention | SNOMED |
| 725898002 | Delirium co-occurrent with dementia | SNOMED |
| 1.42E+14 | Delusions in Alzheimer's disease | SNOMED |
| 421529006 | Dementia associated with AIDS | SNOMED |
| 698781002 | Dementia associated with cerebral anoxia | SNOMED |
| 698624003 | Dementia associated with cerebral lipidosis | SNOMED |
| 698626001 | Dementia associated with multiple sclerosis | SNOMED |
| 698725008 | Dementia associated with neurosyphilis | SNOMED |
| 698625002 | Dementia associated with normal pressure hydrocephalus | SNOMED |
| 425390006 | Dementia associated with Parkinson's Disease | SNOMED |
| 698726009 | Dementia associated with viral encephalitis | SNOMED |
| 733184002 | Dementia caused by heavy metal exposure | SNOMED |
| 722978000 | Dementia caused by toxin | SNOMED |

| | | |
|---|---|---|
| 788898005 | Dementia caused by volatile inhalant | SNOMED |
| 722977005 | Dementia co-occurrent and due to neurocysticercosis | SNOMED |
| 713844000 | Dementia co-occurrent with human immunodeficiency virus infection | SNOMED |
| 762351006 | Dementia due to and following injury of head | SNOMED |
| 722980006 | Dementia due to chromosomal anomaly | SNOMED |
| 733191004 | Dementia due to chronic subdural hematoma | SNOMED |
| 429458009 | Dementia due to Creutzfeldt Jakob disease | SNOMED |
| 724776007 | Dementia due to disorder of central nervous system | SNOMED |
| 733192006 | Dementia due to herpes encephalitis | SNOMED |
| 442344002 | Dementia due to Huntington chorea | SNOMED |
| 724777003 | Dementia due to infectious disease | SNOMED |
| 722979008 | Dementia due to metabolic abnormality | SNOMED |
| 8.24E+13 | Dementia due to multiple sclerosis with altered behavior | SNOMED |
| 1.01E+14 | Dementia due to Parkinson's disease | SNOMED |
| 788899002 | Dementia due to pellagra | SNOMED |
| 2.19E+13 | Dementia due to Pick's disease | SNOMED |
| 733190003 | Dementia due to primary malignant neoplasm of brain | SNOMED |
| 762350007 | Dementia due to prion disease | SNOMED |
| 1.30E+14 | Dementia due to Rett's syndrome | SNOMED |
| 733185001 | Dementia following injury caused by exposure to ionizing radiation | SNOMED |
| 698949001 | Dementia in remission | SNOMED |

| | | |
|---|---|---|
| 278857002 | Dementia of frontal lobe type | SNOMED |
| 1.58E+12 | Dementia of the Alzheimer type with behavioral disturbance | SNOMED |
| 82959004 | Dementia paralytica juvenilis | SNOMED |
| 9.96E+15 | Dementia with aggressive behavior | Nebraska Lexicon |
| 733194007 | Dementia with Down syndrome | SNOMED |
| 733193001 | Dementia with progressive multifocal leukoencephalopathy | SNOMED |
| 1.42E+14 | Depressed mood in Alzheimer's disease | SNOMED |
| 9345005 | Dialysis dementia | SNOMED |
| 288773008 | Difficulty directing attention | SNOMED |
| 1.07E+15 | Difficulty dividing attention | SNOMED |
| 1.07E+15 | Difficulty shifting attention | SNOMED |
| 1.07E+15 | Difficulty sustaining attention | SNOMED |
| 28102002 | Distractibility | SNOMED |
| 416106008 | Disturbance of memory for order of events | SNOMED |
| 288771005 | Does direct attention | SNOMED |
| 1.07E+15 | Does divide attention | SNOMED |
| 288772003 | Does not direct attention | SNOMED |
| 1.07E+15 | Does not divide attention | SNOMED |
| 1.07E+15 | Does not shift attention | SNOMED |
| 1.07E+15 | Does not sustain attention | SNOMED |
| 1.07E+15 | Does shift attention | SNOMED |
| 1.07E+15 | Does sustain attention | SNOMED |
| 1.32E+16 | Early Alzheimers disease | Nebraska Lexicon |
| 1.05E+14 | Early onset Alzheimer's disease with behavioral disturbance | SNOMED |
| 247764002 | Easily distracted | SNOMED |
| 724992007 | Epilepsy co-occurrent and due to dementia | SNOMED |
| 8.24E+13 | Epileptic dementia with behavioral disturbance | SNOMED |
| 247763008 | Excessively focused attention | SNOMED |
| 230265002 | Familial Alzheimer's disease of early onset | SNOMED |

| | | |
|---|---|---|
| 230267005 | Familial Alzheimer's disease of late onset | SNOMED |
| 365080005 | Finding related to ability to direct attention | SNOMED |
| 230269008 | Focal Alzheimer's disease | SNOMED |
| 55533009 | Forgetful | SNOMED |
| 2.42E+12 | Hallucinations co-occurrent and due to late onset dementia | SNOMED |
| 4.81E+14 | High level Alzheimers neuropathology changes | Nebraska Lexicon |
| 283878007 | Impairment of registration | SNOMED |
| 247767009 | Integrated attention for short spells | SNOMED |
| 4.81E+14 | Intermediate level Alzheimers neuropathology changes | Nebraska Lexicon |
| 723123001 | Ischemic vascular dementia | SNOMED |
| 4.71E+14 | Low level Alzheimers neuropathology changes | Nebraska Lexicon |
| 1.67E+16 | Memory deficit due to and following cerebrovascular accident | SNOMED |
| 1.67E+16 | Memory deficit due to and following cerebrovascular disease | SNOMED |
| 1.67E+16 | Memory deficit due to and following hemorrhagic cerebrovascular accident | SNOMED |
| 1.67E+16 | Memory deficit due to and following ischemic cerebrovascular accident | SNOMED |
| 386807006 | Memory impairment | SNOMED |
| 225038006 | Memory lapses | SNOMED |
| 4.28E+14 | Mild dementia | SNOMED |
| 192071009 | Mild memory disturbance | SNOMED |
| 225037001 | Minor memory lapses | SNOMED |
| 230287006 | Mixed cortical and subcortical vascular dementia | SNOMED |
| 7.93E+13 | Mixed dementia | SNOMED |
| 225039003 | Mixes past with present | SNOMED |
| 4.31E+14 | Moderate dementia | SNOMED |

| | | |
|---|---|---|
| 56267009 | Multi-infarct dementia | SNOMED |
| 1.06E+14 | Multi-infarct dementia due to atherosclerosis | SNOMED |
| 722600006 | Non-amnestic Alzheimer disease | SNOMED |
| 230266001 | Non-familial Alzheimer's disease of early onset | SNOMED |
| 230268000 | Non-familial Alzheimer's disease of late onset | SNOMED |
| 86713007 | Nonpersistence | SNOMED |
| 163616009 | O/E - easily distractable | SNOMED |
| 433081000 | Organic amnesia of language | SNOMED |
| 420614009 | Organic dementia associated with AIDS | SNOMED |
| 32541007 | Paramnesia | SNOMED |
| 62239001 | Parkinson-dementia complex of Guam | SNOMED |
| 715737004 | Parkinsonism with dementia of Guadeloupe | SNOMED |
| 230289009 | Patchy dementia | SNOMED |
| 247766000 | Pays attention to own choice of activity | SNOMED |
| 247765001 | Pays fleeting attention | SNOMED |
| 130965009 | Persistence | SNOMED |
| 26329005 | Poor concentration | SNOMED |
| 275277000 | Post-traumatic amnesia | SNOMED |
| 698687007 | Post-traumatic dementia with behavioral change | SNOMED |
| 1.09E+15 | Predominantly cortical dementia | SNOMED |
| 1.09E+15 | Predominantly cortical vascular dementia | SNOMED |
| 421023003 | Presenile dementia associated with AIDS | SNOMED |
| 713488003 | Presenile dementia co-occurrent with human immunodeficiency virus infection | SNOMED |
| 191454001 | Presenile dementia with paranoia | SNOMED |
| 1.09E+15 | Presenile dementia with psychosis | SNOMED |

| | | |
|---|---|---|
| 2.24E+13 | Primary degenerative dementia | SNOMED |
| 698955006 | Primary degenerative dementia of the Alzheimer type, presenile onset in remission | SNOMED |
| 6475002 | Primary degenerative dementia of the Alzheimer type, presenile onset, uncomplicated | SNOMED |
| 65096006 | Primary degenerative dementia of the Alzheimer type, presenile onset, with delirium | SNOMED |
| 54502004 | Primary degenerative dementia of the Alzheimer type, presenile onset, with delusions | SNOMED |
| 10532003 | Primary degenerative dementia of the Alzheimer type, presenile onset, with depression | SNOMED |
| 698954005 | Primary degenerative dementia of the Alzheimer type, senile onset in remission | SNOMED |
| 66108005 | Primary degenerative dementia of the Alzheimer type, senile onset, uncomplicated | SNOMED |
| 4.29E+14 | Primary degenerative dementia of the Alzheimer type, senile onset, with behavioral disturbance | SNOMED |
| 4817008 | Primary degenerative dementia of the Alzheimer type, senile onset, with delirium | SNOMED |
| 55009008 | Primary degenerative dementia of the Alzheimer type, senile onset, with delusions | SNOMED |

| | | |
|---|---|---|
| 26852004 | Primary degenerative dementia of the Alzheimer type, senile onset, with depression | SNOMED |
| 774069007 | PRKAR1B-related neurodegenerative dementia with intermediate filaments | SNOMED |
| 230280008 | Progressive aphasia in Alzheimer's disease | SNOMED |
| 230283005 | Punch drunk syndrome | SNOMED |
| 723390000 | Rapidly progressive dementia | SNOMED |
| 247761005 | Reduced concentration | SNOMED |
| 247762003 | Reduced concentration span | SNOMED |
| 26581009 | Retrospective falsification | SNOMED |
| 425248002 | Scattered attention | SNOMED |
| 25124003 | Selective inattention | SNOMED |
| 15662003 | Senile dementia | SNOMED |
| 312991009 | Senile dementia of the Lewy body type | SNOMED |
| 191457008 | Senile dementia with depressive or paranoid features | SNOMED |
| 191458003 | Senile dementia with paranoia | SNOMED |
| 371026009 | Senile dementia with psychosis | SNOMED |
| 4.28E+14 | Severe dementia | SNOMED |
| 762707000 | Subcortical dementia | SNOMED |
| 90099008 | Subcortical leukoencephalopathy | SNOMED |
| 230286002 | Subcortical vascular dementia | SNOMED |
| 162200009 | Temporary loss of memory | SNOMED |
| 395689002 | Transient epileptic amnesia | SNOMED |
| 307413004 | Transient memory loss | SNOMED |
| 230282000 | Traumatic encephalopathy | SNOMED |
| 60032008 | Unable to concentrate | SNOMED |
| 288770006 | Unable to direct attention | SNOMED |
| 1.07E+15 | Unable to divide attention | SNOMED |
| 1.08E+15 | Unable to shift attention | SNOMED |
| 1.08E+15 | Unable to sustain attention | SNOMED |
| 191463004 | Uncomplicated arteriosclerotic dementia | SNOMED |

| | | |
|---|---|---|
| 698948009 | Vascular dementia in remission | SNOMED |
| 230285003 | Vascular dementia of acute onset | SNOMED |
| 247769007 | Well controlled integrated attention | SNOMED |

Depression

| Code | Name | Vocabulary |
|---|---|---|
| 66344007 | Recurrent major depression | SNOMED |
| 36923009 | Major depression, single episode | SNOMED |
| 13746004 | Bipolar disorder | SNOMED |
| 68890003 | Schizoaffective disorder | SNOMED |
| 36474008 | Severe recurrent major depression without psychotic features | SNOMED |
| 18818009 | Moderate recurrent major depression | SNOMED |
| 191618007 | Bipolar affective disorder, current episode manic | SNOMED |
| 191627008 | Bipolar affective disorder, current episode depression | SNOMED |
| 191613003 | Recurrent major depressive episodes, severe, with psychosis | SNOMED |
| 83225003 | Bipolar II disorder | SNOMED |
| 33135002 | Recurrent major depression in partial remission | SNOMED |
| 16506000 | Mixed bipolar I disorder | SNOMED |
| 76441001 | Severe major depression, single episode, without psychotic features | SNOMED |
| 40379007 | Mild recurrent major depression | SNOMED |
| 371596008 | Bipolar I disorder | SNOMED |
| 191623007 | Bipolar affective disorder, currently manic, severe, with psychosis | SNOMED |
| 38368003 | Schizoaffective disorder, bipolar type | SNOMED |

| | | |
|---|---|---|
| 15639000 | Moderate major depression, single episode | SNOMED |
| 46244001 | Recurrent major depression in full remission | SNOMED |
| 79298009 | Mild major depression, single episode | SNOMED |
| 191611001 | Recurrent major depressive episodes, moderate | SNOMED |
| 19527009 | Single episode of major depression in full remission | SNOMED |
| 68019004 | Recurrent major depression in remission | SNOMED |
| 84760002 | Schizoaffective disorder, depressive type | SNOMED |
| 9340000 | Bipolar I disorder, single manic episode | SNOMED |
| 70747007 | Major depression single episode, in partial remission | SNOMED |
| 61403008 | Severe depressed bipolar I disorder without psychotic features | SNOMED |
| 191570001 | Chronic schizoaffective schizophrenia | SNOMED |
| 162004 | Severe manic bipolar I disorder without psychotic features | SNOMED |
| 191610000 | Recurrent major depressive episodes, mild | SNOMED |
| 191630001 | Bipolar affective disorder, currently depressed, moderate | SNOMED |
| 191641004 | Mixed bipolar affective disorder, severe, with psychosis | SNOMED |
| 191572009 | Acute exacerbation of chronic schizoaffective schizophrenia | SNOMED |
| 31446002 | Bipolar I disorder, most recent episode hypomanic | SNOMED |
| 192362008 | Bipolar affective disorder, current episode mixed | SNOMED |

| | | |
|---|---|---|
| 59617007 | Severe depressed bipolar I disorder with psychotic features | SNOMED |
| 191629006 | Bipolar affective disorder, currently depressed, mild | SNOMED |
| 49512000 | Depressed bipolar I disorder in partial remission | SNOMED |
| 85248005 | Bipolar disorder in remission | SNOMED |
| 765176007 | Psychosis and severe depression co-occurrent and due to bipolar affective disorder | SNOMED |
| 46229002 | Severe mixed bipolar I disorder without psychotic features | SNOMED |
| 191639000 | Mixed bipolar affective disorder, moderate | SNOMED |
| 191621009 | Bipolar affective disorder, currently manic, moderate | SNOMED |
| 63249007 | Manic bipolar I disorder in partial remission | SNOMED |
| 22121000 | Depressed bipolar I disorder in full remission | SNOMED |
| 191638008 | Mixed bipolar affective disorder, mild | SNOMED |
| 41832009 | Severe bipolar I disorder, single manic episode with psychotic features | SNOMED |
| 30935000 | Manic bipolar I disorder in full remission | SNOMED |
| 111485001 | Mixed bipolar I disorder in full remission | SNOMED |
| 191620005 | Bipolar affective disorder, currently manic, mild | SNOMED |
| 36583000 | Mixed bipolar I disorder in partial remission | SNOMED |
| 191569002 | Subchronic schizoaffective schizophrenia | SNOMED |
| 191616006 | Recurrent depression | SNOMED |
| 5703000 | Bipolar disorder in partial remission | SNOMED |

| | | |
|---|---|---|
| 14495005 | Severe bipolar I disorder, single manic episode without psychotic features | SNOMED |
| 41836007 | Bipolar disorder in full remission | SNOMED |
| 191571002 | Acute exacerbation of subchronic schizoaffective schizophrenia | SNOMED |
| 41552001 | Mild bipolar I disorder, single manic episode | SNOMED |
| 191634005 | Bipolar affective disorder, currently depressed, in full remission | SNOMED |
| 28884001 | Moderate bipolar I disorder, single manic episode | SNOMED |
| 191625000 | Bipolar affective disorder, currently manic, in full remission | SNOMED |
| 191574005 | Schizoaffective schizophrenia in remission | SNOMED |
| 191643001 | Mixed bipolar affective disorder, in full remission | SNOMED |
| 75360000 | Bipolar I disorder, single manic episode, in remission | SNOMED |
| 3530005 | Bipolar I disorder, single manic episode, in full remission | SNOMED |
| 7.55E+14 | [X]Recurrent major depressive episodes, severe, with psychosis, psychosis in remission | SNOMED |
| 7.55E+14 | [X]Single major depressive episode, severe, with psychosis, psychosis in remission | SNOMED |
| 767633005 | Bipolar affective disorder, most recent episode mixed | SNOMED |
| 1.45E+16 | Bipolar disease in pregnancy | Nebraska Lexicon |
| 1.62E+16 | Bipolar disorder caused by drug | SNOMED |
| 767631007 | Bipolar disorder, most recent episode depression | SNOMED |

| | | |
|---|---|---|
| 767632000 | Bipolar disorder, most recent episode manic | SNOMED |
| 29929003 | Bipolar I disorder, most recent episode depressed with atypical features | SNOMED |
| 21900002 | Bipolar I disorder, most recent episode depressed with catatonic features | SNOMED |
| 75752004 | Bipolar I disorder, most recent episode depressed with melancholic features | SNOMED |
| 87203005 | Bipolar I disorder, most recent episode depressed with postpartum onset | SNOMED |
| 767636002 | Bipolar I disorder, most recent episode depression | SNOMED |
| 767635003 | Bipolar I disorder, most recent episode manic | SNOMED |
| 17782008 | Bipolar I disorder, most recent episode manic with catatonic features | SNOMED |
| 55516002 | Bipolar I disorder, most recent episode manic with postpartum onset | SNOMED |
| 73471000 | Bipolar I disorder, most recent episode mixed with catatonic features | SNOMED |
| 65042007 | Bipolar I disorder, most recent episode mixed with postpartum onset | SNOMED |
| 87950005 | Bipolar I disorder, single manic episode with catatonic features | SNOMED |
| 1499003 | Bipolar I disorder, single manic episode with postpartum onset | SNOMED |
| 78269000 | Bipolar I disorder, single manic episode, in partial remission | SNOMED |
| 48937005 | Bipolar II disorder, most recent episode hypomanic | SNOMED |

| | | |
|---|---|---|
| 16295005 | Bipolar II disorder, most recent episode major depressive | SNOMED |
| 43568002 | Bipolar II disorder, most recent episode major depressive with atypical features | SNOMED |
| 22407005 | Bipolar II disorder, most recent episode major depressive with catatonic features | SNOMED |
| 34315001 | Bipolar II disorder, most recent episode major depressive with melancholic features | SNOMED |
| 30687003 | Bipolar II disorder, most recent episode major depressive with postpartum onset | SNOMED |
| 723903001 | Bipolar type I disorder currently in full remission | SNOMED |
| 723905008 | Bipolar type II disorder currently in full remission | SNOMED |
| 51637008 | Chronic bipolar I disorder, most recent episode depressed | SNOMED |
| 1196001 | Chronic bipolar II disorder, most recent episode major depressive | SNOMED |
| 14183003 | Chronic major depressive disorder, single episode | SNOMED |
| 2618002 | Chronic recurrent major depressive disorder | SNOMED |
| 49468007 | Depressed bipolar I disorder | SNOMED |
| 53607008 | Depressed bipolar I disorder in remission | SNOMED |
| 274948002 | Endogenous depression - recurrent | SNOMED |
| 42925002 | Major depressive disorder, single episode with atypical features | SNOMED |

| | | |
|---|---|---|
| 69392006 | Major depressive disorder, single episode with catatonic features | SNOMED |
| 63778009 | Major depressive disorder, single episode with melancholic features | SNOMED |
| 25922000 | Major depressive disorder, single episode with postpartum onset | SNOMED |
| 68569003 | Manic bipolar I disorder | SNOMED |
| 45479006 | Manic bipolar I disorder in remission | SNOMED |
| 13313007 | Mild bipolar disorder | SNOMED |
| 71294008 | Mild bipolar II disorder, most recent episode major depressive | SNOMED |
| 74686005 | Mild depressed bipolar I disorder | SNOMED |
| 1.63E+16 | Mild major depressive disorder co-occurrent with anxiety single episode | SNOMED |
| 71984005 | Mild manic bipolar I disorder | SNOMED |
| 43769008 | Mild mixed bipolar I disorder | SNOMED |
| 720454007 | Minimal major depression single episode | SNOMED |
| 720451004 | Minimal recurrent major depression | SNOMED |
| 191636007 | Mixed bipolar affective disorder | SNOMED |
| 7.61E+14 | Mixed bipolar affective disorder, in partial remission | SNOMED |
| 7.65E+14 | Mixed bipolar affective disorder, severe | SNOMED |
| 35481005 | Mixed bipolar I disorder in remission | SNOMED |
| 79584002 | Moderate bipolar disorder | SNOMED |
| 35846004 | Moderate bipolar II disorder, most recent episode major depressive | SNOMED |
| 66631006 | Moderate depressed bipolar I disorder | SNOMED |

| | | |
|---|---|---|
| 1.63E+16 | Moderate major depressive disorder co-occurrent with anxiety single episode | SNOMED |
| 82998009 | Moderate manic bipolar I disorder | SNOMED |
| 40926005 | Moderate mixed bipolar I disorder | SNOMED |
| 720453001 | Moderately severe major depression single episode | SNOMED |
| 720452006 | Moderately severe recurrent major depression | SNOMED |
| 231444002 | Organic bipolar disorder | SNOMED |
| 1.33E+14 | Rapid cycling bipolar I disorder | SNOMED |
| 789061003 | Rapid cycling bipolar II disorder | SNOMED |
| 40568001 | Recurrent brief depressive disorder | SNOMED |
| 1.09E+15 | Recurrent depression with current moderate episode | SNOMED |
| 1.09E+15 | Recurrent depression with current severe episode and psychotic features | SNOMED |
| 1.09E+15 | Recurrent depression with current severe episode without psychotic features | SNOMED |
| 1.63E+16 | Recurrent major depressive disorder co-occurrent with anxiety in full remission | SNOMED |
| 1.63E+16 | Recurrent major depressive disorder in partial remission co-occurrent with anxiety | SNOMED |
| 38694004 | Recurrent major depressive disorder with atypical features | SNOMED |
| 39809009 | Recurrent major depressive disorder with catatonic features | SNOMED |
| 319768000 | Recurrent major depressive disorder with melancholic features | SNOMED |

| | | |
|---|---|---|
| 71336009 | Recurrent major depressive disorder with postpartum onset | SNOMED |
| 268621008 | Recurrent major depressive episodes | SNOMED |
| 7.65E+14 | Recurrent major depressive episodes, in partial remission | SNOMED |
| 7.65E+14 | Recurrent major depressive episodes, in remission | SNOMED |
| 7.65E+14 | Recurrent major depressive episodes, severe | SNOMED |
| 1.63E+16 | Recurrent mild major depressive disorder co-occurrent with anxiety | SNOMED |
| 1.63E+16 | Recurrent moderate major depressive disorder co-occurrent with anxiety | SNOMED |
| 1.63E+16 | Recurrent severe major depressive disorder co-occurrent with anxiety | SNOMED |
| 271428004 | Schizoaffective disorder, manic type | SNOMED |
| 270901009 | Schizoaffective disorder, mixed type | SNOMED |
| 191567000 | Schizoaffective schizophrenia | SNOMED |
| 247803002 | Seasonal affective disorder | SNOMED |
| 371600003 | Severe bipolar disorder | SNOMED |
| 4441000 | Severe bipolar disorder with psychotic features | SNOMED |
| 70546001 | Severe bipolar disorder with psychotic features, mood-congruent | SNOMED |
| 26530004 | Severe bipolar disorder with psychotic features, mood-incongruent | SNOMED |
| 53049002 | Severe bipolar disorder without psychotic features | SNOMED |
| 371599001 | Severe bipolar I disorder | SNOMED |
| 13581000 | Severe bipolar I disorder, single manic episode with psychotic features, mood-congruent | SNOMED |

| | | |
|---|---|---|
| 86058007 | Severe bipolar I disorder, single manic episode with psychotic features, mood-incongruent | SNOMED |
| 371604007 | Severe bipolar II disorder | SNOMED |
| 30520009 | Severe bipolar II disorder, most recent episode major depressive with psychotic features | SNOMED |
| 19300006 | Severe bipolar II disorder, most recent episode major depressive with psychotic features, mood-congruent | SNOMED |
| 20960007 | Severe bipolar II disorder, most recent episode major depressive with psychotic features, mood-incongruent | SNOMED |
| 81319007 | Severe bipolar II disorder, most recent episode major depressive without psychotic features | SNOMED |
| 12969000 | Severe bipolar II disorder, most recent episode major depressive, in full remission | SNOMED |
| 67002003 | Severe bipolar II disorder, most recent episode major depressive, in partial remission | SNOMED |
| 35722002 | Severe bipolar II disorder, most recent episode major depressive, in remission | SNOMED |
| 2.61E+11 | Severe depressed bipolar I disorder | SNOMED |
| 54761006 | Severe depressed bipolar I disorder with psychotic features, mood-congruent | SNOMED |
| 26203008 | Severe depressed bipolar I disorder with psychotic features, mood-incongruent | SNOMED |
| 2.51E+11 | Severe major depression, single episode | SNOMED |
| 77911002 | Severe major depression, single episode, with | SNOMED |

| | | |
|---|---|---|
| | psychotic features, mood-congruent | |
| 20250007 | Severe major depression, single episode, with psychotic features, mood-incongruent | SNOMED |
| 1.63E+16 | Severe major depressive disorder co-occurrent with anxiety single episode | SNOMED |
| 2.37E+13 | Severe manic bipolar I disorder | SNOMED |
| 28663008 | Severe manic bipolar I disorder with psychotic features | SNOMED |
| 78640000 | Severe manic bipolar I disorder with psychotic features, mood-congruent | SNOMED |
| 33380008 | Severe manic bipolar I disorder with psychotic features, mood-incongruent | SNOMED |
| 2.71E+11 | Severe mixed bipolar I disorder | SNOMED |
| 10981006 | Severe mixed bipolar I disorder with psychotic features | SNOMED |
| 64731001 | Severe mixed bipolar I disorder with psychotic features, mood-congruent | SNOMED |
| 10875004 | Severe mixed bipolar I disorder with psychotic features, mood-incongruent | SNOMED |
| 2.81E+11 | Severe recurrent major depression | SNOMED |
| 28475009 | Severe recurrent major depression with psychotic features | SNOMED |
| 33078009 | Severe recurrent major depression with psychotic features, mood-congruent | SNOMED |
| 15193003 | Severe recurrent major depression with psychotic features, mood-incongruent | SNOMED |

| | | |
|---|---|---|
| 1.33E+14 | Severe seasonal affective disorder | SNOMED |
| 7.65E+14 | Single major depressive episode, in remission | SNOMED |
| 191604000 | Single major depressive episode, severe, with psychosis | SNOMED |

Dyspnea

| Code | Name | Vocabulary |
|---|---|---|
| 267036007 | Dyspnea | SNOMED |
| 60845006 | Dyspnea on exertion | SNOMED |
| 397702004 | Borg Breathlessness Score finding | SNOMED |
| 401275008 | Borg Breathlessness Score: 0 none at all | SNOMED |
| 401323002 | Borg Breathlessness Score: 0.5 very, very slight | SNOMED |
| 401279002 | Borg Breathlessness Score: 1 very slight | SNOMED |
| 401293009 | Borg Breathlessness Score: 10 maximal | SNOMED |
| 401280004 | Borg Breathlessness Score: 2 slight | SNOMED |
| 401281000 | Borg Breathlessness Score: 3 moderate | SNOMED |
| 401282007 | Borg Breathlessness Score: 4 somewhat severe | SNOMED |
| 401284008 | Borg Breathlessness Score: 5 severe | SNOMED |
| 401286005 | Borg Breathlessness Score: 6 severe | SNOMED |
| 401290007 | Borg Breathlessness Score: 7 very severe | SNOMED |
| 401291006 | Borg Breathlessness Score: 8 very severe | SNOMED |
| 401292004 | Borg Breathlessness Score: 9 very, very severe | SNOMED |
| 161940008 | Breathless - mild exertion | SNOMED |
| 161939006 | Breathless - moderate exertion | SNOMED |

| | | |
|---|---|---|
| 390871002 | Breathless - strenuous exertion | SNOMED |
| 57769004 | Dyspnea after eating | SNOMED |
| 422177004 | Dyspnea associated with AIDS | SNOMED |
| 161941007 | Dyspnea at rest | SNOMED |
| 20112008 | Dyspnea leaning over | SNOMED |
| 24921003 | Dyspnea raising arms | SNOMED |
| 17216000 | Dyspnea, class I | SNOMED |
| 72365000 | Dyspnea, class II | SNOMED |
| 39950000 | Dyspnea, class III | SNOMED |
| 73322006 | Dyspnea, class IV | SNOMED |
| 1.09E+15 | eMRC (extended Medical Research Council) dyspnoea scale grade 1 | SNOMED |
| 1.09E+15 | eMRC (extended Medical Research Council) dyspnoea scale grade 2 | SNOMED |
| 1.09E+15 | eMRC (extended Medical Research Council) dyspnoea scale grade 3 | SNOMED |
| 1.09E+15 | eMRC (extended Medical Research Council) dyspnoea scale grade 4 | SNOMED |
| 1.09E+15 | eMRC (extended Medical Research Council) dyspnoea scale grade 5a | SNOMED |
| 1.09E+15 | eMRC (extended Medical Research Council) dyspnoea scale grade 5b | SNOMED |
| 34560001 | Expiratory dyspnea | SNOMED |
| 23141003 | Gasping for breath | SNOMED |
| 297216006 | Increasing breathlessness | SNOMED |
| 25209001 | Inspiratory dyspnea | SNOMED |
| 391120009 | Medical Research Council Dyspnoea scale grade 1 | SNOMED |
| 391123006 | Medical Research Council Dyspnoea scale grade 2 | SNOMED |
| 391124000 | Medical Research Council Dyspnoea scale grade 3 | SNOMED |
| 391125004 | Medical Research Council Dyspnoea scale grade 4 | SNOMED |

| | | |
|---|---|---|
| 391126003 | Medical Research Council Dyspnoea scale grade 5 | SNOMED |
| 8.52E+14 | Minimal breathlessness | SNOMED |
| 1.10E+15 | mMRC (modified Medical Research Council) dyspnoea scale grade 0 | SNOMED |
| 1.10E+15 | mMRC (modified Medical Research Council) dyspnoea scale grade 1 | SNOMED |
| 1.10E+15 | mMRC (modified Medical Research Council) dyspnoea scale grade 2 | SNOMED |
| 1.10E+15 | mMRC (modified Medical Research Council) dyspnoea scale grade 3 | SNOMED |
| 1.10E+15 | mMRC (modified Medical Research Council) dyspnoea scale grade 4 | SNOMED |
| 248548009 | Nocturnal dyspnea | SNOMED |
| 162890008 | O/E - dyspnea | SNOMED |
| 59265000 | Paroxysmal dyspnea | SNOMED |
| 55442000 | Paroxysmal nocturnal dyspnea | SNOMED |
| 30744009 | Platypnea | SNOMED |
| 390870001 | Short of breath dressing/undressing | SNOMED |
| 102577000 | Trepopnea | SNOMED |
| 407588003 | Unable to complete a sentence in one breath | SNOMED |

Fever

| Code | Name | Vocabulary |
|---|---|---|
| 386661006 | Fever | SNOMED |
| 41497008 | Febrile convulsion | SNOMED |
| 432354000 | Simple febrile seizure | SNOMED |
| 420079008 | Relapsing fever | SNOMED |
| 433083002 | Complex febrile seizure | SNOMED |
| 722892007 | Fever due to infection | SNOMED |
| 186694006 | Sweating fever | SNOMED |
| 1.02E+13 | Viral fever | SNOMED |
| 704425001 | Chronic fever | SNOMED |
| 7520000 | Pyrexia of unknown origin | SNOMED |

| | | |
|---|---|---|
| 9619006 | Aseptic fever | SNOMED |
| 77957000 | Intermittent fever | SNOMED |
| 63993003 | Remittent fever | SNOMED |
| 365989009 | Phase of fever - finding | SNOMED |
| 365977007 | Pattern of fever - finding | SNOMED |
| 409702008 | Hyperpyrexia | SNOMED |
| 42136008 | Swinging fever | SNOMED |
| 274640006 | Fever with chills | SNOMED |
| 271753002 | Irregular fever | SNOMED |
| 271752007 | Staircase fever | SNOMED |
| 271751000 | Continuous fever | SNOMED |
| 271750004 | Gradual rise of fever | SNOMED |
| 271749004 | Acute rise of fever | SNOMED |
| 271897009 | O/E - fever | SNOMED |
| 271755009 | Gradual fall of fever | SNOMED |
| 271754008 | Rapid fall of fever | SNOMED |
| 307200007 | Recurrent febrile convulsion | SNOMED |
| 426000000 | Fever greater than 100.4 Fahrenheit | SNOMED |
| 304213008 | Low grade pyrexia | SNOMED |
| 248447001 | Fever defervescence | SNOMED |
| 248443002 | Intermittent hepatic fever | SNOMED |
| 248436008 | Slightly remittent fever | SNOMED |
| 248432005 | Rising phase of fever | SNOMED |
| 248435007 | Prolonged fever | SNOMED |
| 248449003 | Central fever | SNOMED |
| 248445009 | Fever, diurnal variation | SNOMED |
| 248444008 | Biphasic fever | SNOMED |
| 248434006 | Falling phase of fever | SNOMED |
| 248433000 | Plateau phase of fever | SNOMED |
| 135883003 | Cough with fever | SNOMED |
| 230432008 | Familial febrile convulsions | SNOMED |
| 164311002 | O/E - fever - remittent | SNOMED |
| 164316007 | O/E - fever-gradual fall-lysis | SNOMED |
| 164314005 | O/E - fever - irregular | SNOMED |
| 164309006 | O/E - fever - continuous | SNOMED |
| 164315006 | O/E - fever - fast fall-crisis | SNOMED |
| 164307008 | O/E - fever - acute rise | SNOMED |
| 164288004 | O/E - pyrexia of unknown origin | SNOMED |
| 164312009 | O/E - fever - intermittent | SNOMED |
| 164308003 | O/E - fever - gradual rise | SNOMED |

| | | |
|---|---|---|
| 164304001 | O/E - hyperpyrexia - greater than 40.5 degrees Celsius | SNOMED |
| 163595003 | O/E - febrile convulsion | SNOMED |
| 102496004 | Spiking fever | SNOMED |
| 111950007 | Factitious fever | SNOMED |
| 1.14E+16 | Neutropenic fever | Nebraska Lexicon |
| 4.34E+14 | Complex febrile seizure, refractory | SNOMED |
| 4.34E+14 | Simple febrile seizure, non-refractory | SNOMED |
| 4.34E+14 | Simple febrile seizure, refractory | SNOMED |
| 4.34E+14 | Complex febrile seizure, non-refractory | SNOMED |

Influenza Conditions

| Code | Name | Vocabulary |
|---|---|---|
| 1.07E+16 | Upper respiratory tract infection due to Influenza | SNOMED |
| 195878008 | Pneumonia and influenza | SNOMED |
| 772839003 | Pneumonia caused by Influenza A virus | SNOMED |
| 6142004 | Influenza | SNOMED |
| 3.29E+14 | Upper respiratory tract infection due to Influenza A | SNOMED |
| 61700007 | Influenza with non-respiratory manifestation | SNOMED |
| 1.43E+14 | Upper respiratory tract infection due to H1N1 influenza | SNOMED |
| 442696006 | Influenza due to Influenza A virus subtype H1N1 | SNOMED |
| 1.43E+14 | Pneumonia due to H1N1 influenza | SNOMED |
| 772828001 | Influenza caused by Influenza A virus subtype H5N1 | SNOMED |
| 442438000 | Influenza due to Influenza A virus | SNOMED |
| 195929004 | Influenza with gastrointestinal tract involvement | SNOMED |
| 1.06E+16 | Otitis media due to influenza | SNOMED |

| | | |
|---|---|---|
| 74644004 | Influenza with encephalopathy | SNOMED |
| 1.07E+16 | Encephalopathy due to Influenza A virus | SNOMED |
| 1.43E+14 | Gastroenteritis due to H1N1 influenza | SNOMED |
| 1.63E+13 | Pneumonia due to influenza | SNOMED |
| 1.43E+14 | Encephalopathy due to H1N1 influenza | SNOMED |
| 1.43E+14 | Myocarditis due to Influenza A virus subtype H1N1 | SNOMED |
| 1.07E+16 | Otitis media due to Influenza A virus | SNOMED |
| 1.06E+16 | Myocarditis due to Influenza A virus | SNOMED |
| 1.06E+16 | Bronchiolitis caused by influenza virus | SNOMED |
| 1.06E+16 | Gastroenteritis due to Influenza A virus | SNOMED |
| 1.06E+16 | Gastroenteritis due to influenza | SNOMED |
| 1.06E+16 | Otitis media due to H1N1 influenza | SNOMED |
| 707448003 | Influenza due to Influenza A virus subtype H7N9 | SNOMED |
| 450716003 | Influenza due to Influenza A virus subtype H9 | SNOMED |
| 450715004 | Influenza due to Influenza A virus subtype H7 | SNOMED |
| 738276008 | Influenza with CNS disorder | SNOMED |
| 1.03E+15 | Influenza with pneumonia due to seasonal influenza virus | SNOMED |
| 1.03E+15 | Influenza due to seasonal influenza virus | SNOMED |
| 1.03E+15 | Influenza due to pandemic influenza virus | SNOMED |
| 1.03E+15 | Influenza due to zoonotic influenza virus | SNOMED |
| 713083002 | Influenza caused by Influenza A virus subtype H5 | SNOMED |
| 719865001 | Influenza caused by pandemic influenza virus | SNOMED |

| | | |
|---|---|---|
| 719590007 | Influenza caused by seasonal influenza virus | SNOMED |
| 772810003 | Influenza caused by Influenza A virus subtype H3N2 | SNOMED |
| 81524006 | Influenza due to Influenza C virus | SNOMED |
| 78046005 | Myocarditis due to influenza virus | SNOMED |
| 408687004 | Healthcare associated influenza disease | SNOMED |
| 41269000 | Influenzal bronchopneumonia | SNOMED |
| 43692000 | Influenzal acute upper respiratory infection | SNOMED |
| 309789002 | Encephalitis due to influenza | SNOMED |
| 195923003 | Influenza with laryngitis | SNOMED |
| 195924009 | Influenza with pharyngitis | SNOMED |
| 194946005 | Acute myocarditis - influenzal | SNOMED |
| 24662006 | Influenza due to Influenza B virus | SNOMED |
| 4.35E+14 | Influenza due to Influenza A virus with upper respiratory signs | SNOMED |
| 4.35E+14 | Pneumonia due to Influenza A virus subtype H1N1 | SNOMED |
| 4.35E+14 | Pneumonia due to Influenza A virus | SNOMED |

Precoordinated Influenza Conditions

| Code | Name | Vocabulary |
|---|---|---|
| 702482001 | Influenza A H1N1 virus 2009 pandemic strain present | SNOMED |
| 1.81E+11 | Influenza A virus present | SNOMED |
| 2.31E+11 | Influenza A virus subtype H1 2009 pandemic strain present | SNOMED |
| 441043003 | Influenza A virus subtype H1 present | SNOMED |
| 711330007 | Influenza A virus subtype H1N1 detected | SNOMED |

| | | |
|---|---|---|
| 440927002 | Influenza A virus subtype H2 present | SNOMED |
| 441049004 | Influenza A virus subtype H3 present | SNOMED |
| 1.01E+12 | Influenza A virus subtype H5 asian strain detected | SNOMED |
| 441343005 | Influenza A virus subtype H5 present | SNOMED |
| 708119004 | Influenza A virus subtype H7 present | SNOMED |
| 708120005 | Influenza A virus subtype H9 present | SNOMED |
| 699872005 | Influenza A virus untyped strain present | SNOMED |
| 441345003 | Influenza B virus present | SNOMED |

Influenza Tests

| Code | Name | Vocabulary |
|---|---|---|
| 49520-0 | Influenza virus A H1 RNA [Presence] in Isolate by NAA with probe detection | LOINC |
| 49535-8 | Influenza virus B RNA [Presence] in Isolate by NAA with probe detection | LOINC |
| 49531-7 | Influenza virus A RNA [Presence] in Isolate by NAA with probe detection | LOINC |
| 49523-4 | Influenza virus A H3 RNA [Presence] in Isolate by NAA with probe detection | LOINC |
| 31437-7 | Influenza virus A IgG Ab [Units/volume] in Serum | LOINC |
| 7920-2 | Influenza virus A Ab [Units/volume] in Serum | LOINC |
| 22365-1 | Influenza virus A Ab [Titer] in Serum | LOINC |

Joint Pain

| Code | Name | Vocabulary |
|---|---|---|
| 267949000 | Shoulder joint pain | SNOMED |

| | | |
|---|---|---|
| 30989003 | Knee pain | SNOMED |
| 267954009 | Arthralgia of the ankle and/or foot | SNOMED |
| 57676002 | Joint pain | SNOMED |
| 49218002 | Hip pain | SNOMED |
| 713413001 | Joint pain of pelvic region | SNOMED |
| 3.17E+14 | Pain in right knee | SNOMED |
| 3.17E+14 | Pain in left knee | SNOMED |
| 202482009 | Wrist joint pain | SNOMED |
| 202480001 | Elbow joint pain | SNOMED |
| 202472008 | Hand joint pain | SNOMED |
| 35678005 | Multiple joint pain | SNOMED |
| 91943004 | Arthralgia of temporomandibular joint | SNOMED |
| 202479004 | Acromioclavicular joint pain | SNOMED |
| 1.13E+14 | Acute arthralgia of knee | SNOMED |
| 299447008 | Ankle joint - painful on movement | SNOMED |
| 202490009 | Ankle joint pain | SNOMED |
| 239733006 | Anterior knee pain | SNOMED |
| 772826002 | Anti-cyclic citrullinated peptide antibody positive arthralgia | SNOMED |
| 1.09E+15 | Anti-cyclic citrullinated peptide antibody positive arthralgia | SNOMED |
| 1.59E+16 | Arthralgia assoc with cystic fibrosis | Nebraska Lexicon |
| 267952008 | Arthralgia of the pelvic region and thigh | SNOMED |
| 267950000 | Arthralgia of the upper arm | SNOMED |
| 1.56E+16 | Bilateral acromioclavicular joint pain | SNOMED |
| 1.22E+16 | Bilateral ankle joint pain | SNOMED |
| 1.24E+16 | Bilateral chronic pain following total hip arthroplasty | SNOMED |
| 1.22E+16 | Bilateral elbow joint pain | SNOMED |
| 1.22E+16 | Bilateral foot joint pain | SNOMED |
| 1.22E+16 | Bilateral hip joint pain | SNOMED |
| 1.19E+16 | Bilateral knee pain | SNOMED |
| 1.69E+16 | Bilateral pain of joint of hands | SNOMED |

| | | |
|---|---|---|
| 1.56E+16 | Bilateral sacroiliac joint pain | SNOMED |
| 1.22E+16 | Bilateral shoulder joint pain | SNOMED |
| 1.56E+16 | Bilateral temporomandibular joint pain | SNOMED |
| 1.59E+16 | Bilateral tenderness of temporomandibular joints | SNOMED |
| 1.24E+16 | Bilateral total knee chronic pain following arthroplasty | SNOMED |
| 298253002 | Cervical facet joint pain | SNOMED |
| 298478001 | Cervical spine painful on movement | SNOMED |
| 736428003 | Chronic arthralgia of temporomandibular joint | SNOMED |
| 1.19E+16 | Chronic arthralgias of hips and knees | Nebraska Lexicon |
| 1.24E+16 | Chronic pain following left total hip arthroplasty | SNOMED |
| 1.24E+16 | Chronic pain following left total knee arthroplasty | SNOMED |
| 1.24E+16 | Chronic pain following right total hip arthroplasty | SNOMED |
| 1.24E+16 | Chronic pain following right total knee arthroplasty | SNOMED |
| 782661001 | Chronic sacroiliac joint pain | SNOMED |
| 1.09E+15 | Chronic sacroiliac joint pain | SNOMED |
| 7.58E+15 | Costal chondral thoracic pain | Nebraska Lexicon |
| 202485006 | Distal interphalangeal joint of finger pain | SNOMED |
| 202481002 | Distal radioulnar joint pain | SNOMED |
| 298929004 | Elbow joint - painful on movement | SNOMED |
| 247369005 | Facet joint pain | SNOMED |
| 299112005 | Finger joint painful on movement | SNOMED |
| 202495004 | First metatarsophalangeal joint pain | SNOMED |
| 299513007 | Foot joint - painful on movement | SNOMED |
| 279066007 | Foot joint pain | SNOMED |
| 2.72E+16 | Forearm joint pain | Nebraska Lexicon |
| 4.49E+14 | Herniation of lumbar intervertebral disc with sciatica | SNOMED |

| | | |
|---|---|---|
| 299308007 | Hip joint painful on movement | SNOMED |
| 736447000 | Infrequent episodic arthralgia of temporomandibular joint | SNOMED |
| 202497007 | Interphalangeal joint of toe pain | SNOMED |
| 1.08E+15 | Joint pain in left hand | SNOMED |
| 1.08E+15 | Joint pain in right hand | SNOMED |
| 299377003 | Knee joint painful on movement | SNOMED |
| 202496003 | Lesser metatarsophalangeal joint pain | SNOMED |
| 46960006 | Lumbago-sciatica due to displacement of lumbar intervertebral disc | SNOMED |
| 279063004 | Lumbar facet joint pain | SNOMED |
| 298674008 | Lumbar spine painful on movement | SNOMED |
| 202483004 | Metacarpophalangeal joint pain | SNOMED |
| 279067003 | Metatarsophalangeal joint pain | SNOMED |
| 164539000 | O/E - joint movement painful | SNOMED |
| 309765009 | Pain due to hip joint prosthesis | SNOMED |
| 309767001 | Pain due to knee joint prosthesis | SNOMED |
| 309768006 | Pain due to shoulder joint prosthesis | SNOMED |
| 1.22E+16 | Pain in left sacroiliac joint | SNOMED |
| 3.17E+14 | Pain in right hip joint | SNOMED |
| 1.22E+16 | Pain in right sacroiliac joint | SNOMED |
| 433017009 | Pain in symphysis pubis in pregnancy | SNOMED |
| 1.57E+16 | Pain of bilateral sternoclavicular joints | SNOMED |
| 1.57E+16 | Pain of joint of bilateral lower legs | SNOMED |
| 1.08E+15 | Pain of joint of left foot | SNOMED |
| 1.57E+16 | Pain of joint of left lower leg | SNOMED |
| 1.08E+15 | Pain of joint of right foot | SNOMED |
| 1.57E+16 | Pain of joint of right lower leg | SNOMED |

| | | |
|---|---|---|
| 774137000 | Pain of left acromioclavicular joint | SNOMED |
| 1.59E+16 | Pain of left ankle joint | SNOMED |
| 1.59E+16 | Pain of left elbow joint | SNOMED |
| 3.17E+14 | Pain of left hip joint | SNOMED |
| 1.59E+16 | Pain of left shoulder joint | SNOMED |
| 1.08E+15 | Pain of left sternoclavicular joint | SNOMED |
| 1.22E+16 | Pain of left temporomandibular joint | SNOMED |
| 774136009 | Pain of right acromioclavicular joint | SNOMED |
| 1.59E+16 | Pain of right ankle joint | SNOMED |
| 1.59E+16 | Pain of right elbow joint | SNOMED |
| 1.59E+16 | Pain of right shoulder joint | SNOMED |
| 1.08E+15 | Pain of right sternoclavicular joint | SNOMED |
| 1.22E+16 | Pain of right temporomandibular joint | SNOMED |
| 298255009 | Pain on joint movement | SNOMED |
| 429531000 | Pain on passive stretch of joint | SNOMED |
| 387638003 | Painful swelling of joint | SNOMED |
| 1.22E+16 | Patellofemoral syndrome of bilateral knees | SNOMED |
| 1.08E+15 | Patellofemoral syndrome of left knee | SNOMED |
| 1.08E+15 | Patellofemoral syndrome of right knee | SNOMED |
| 703619001 | Pelvic girdle pain | SNOMED |
| 240271006 | Persistent prosthetic joint pain | SNOMED |
| 202484005 | Proximal interphalangeal joint of finger pain | SNOMED |
| 277138006 | Rubella arthralgia | SNOMED |
| 202487003 | Sacroiliac joint pain | SNOMED |
| 298858000 | Shoulder joint - painful arc | SNOMED |
| 9.21E+14 | Shoulder joint painful on external rotation | SNOMED |
| 298857005 | Shoulder joint painful on movement | SNOMED |
| 202478007 | Sternoclavicular joint pain | SNOMED |
| 202491008 | Subtalar joint pain | SNOMED |

| | | |
|---|---|---|
| 299554004 | Subtalar joint painful on movement | SNOMED |
| 202493006 | Talonavicular joint pain | SNOMED |
| 298376001 | Temporomandibular joint painful on movement | SNOMED |
| 299446004 | Tenderness of ankle joint | SNOMED |
| 298928007 | Tenderness of elbow joint | SNOMED |
| 299111003 | Tenderness of finger joint | SNOMED |
| 299512002 | Tenderness of foot joint | SNOMED |
| 299307002 | Tenderness of hip joint | SNOMED |
| 110288007 | Tenderness of joint | SNOMED |
| 299372009 | Tenderness of knee joint | SNOMED |
| 1.22E+16 | Tenderness of left temporomandibular joint | SNOMED |
| 1.22E+16 | Tenderness of right temporomandibular joint | SNOMED |
| 298251000 | Tenderness of sacroiliac joint | SNOMED |
| 298856001 | Tenderness of shoulder joint | SNOMED |
| 299553005 | Tenderness of subtalar joint | SNOMED |
| 298375002 | Tenderness of temporomandibular joint | SNOMED |
| 109659002 | Tenderness of temporomandibular joint on palpation | SNOMED |
| 299198008 | Tenderness of thumb joint | SNOMED |
| 299632005 | Tenderness of toe joint | SNOMED |
| 299017002 | Tenderness of wrist joint | SNOMED |
| 298254008 | Thoracic facet joint pain | SNOMED |
| 298579007 | Thoracic spine - painful on movement | SNOMED |
| 299199000 | Thumb joint painful on movement | SNOMED |
| 202489000 | Tibiofibular joint pain | SNOMED |
| 299633000 | Toe joint painful on movement | SNOMED |
| 8.74E+13 | Vertebral joint pain | SNOMED |
| 299018007 | Wrist joint painful on movement | SNOMED |

Lung Disorder
Any descendant of SNOMED-CT 19829001 (“Disorder of Lung”).

Malaise or Fatigue

| Code | Name | Vocabulary |
|---|---|---|
| 367391008 | Malaise | SNOMED |
| 84229001 | Fatigue | SNOMED |
| 780.79 | Other malaise and fatigue | ICD9CM |
| 13791008 | Asthenia | SNOMED |
| 52702003 | Chronic fatigue syndrome | SNOMED |
| 271795006 | Malaise and fatigue | SNOMED |
| 780.7 | Malaise and fatigue | ICD9CM |
| 1.13E+12 | Asthenia due to disease | SNOMED |
| 248278004 | Attacks of weakness | SNOMED |
| 1.56E+16 | Bilateral weakness of upper limbs | SNOMED |
| 161874006 | Heavy feeling | SNOMED |
| 161873000 | Heavy legs | SNOMED |
| R53 | Malaise and fatigue | ICD10CM |
| 3.77E+14 | Mild chronic fatigue syndrome | SNOMED |
| 3.77E+14 | Moderate chronic fatigue syndrome | SNOMED |
| 713568000 | Occasionally tired | SNOMED |
| R53.8 | Other malaise and fatigue | ICD10CM |
| 442099003 | Psychogenic fatigue | SNOMED |
| 373931001 | Sensation of heaviness in limbs | SNOMED |
| 3.77E+14 | Severe chronic fatigue syndrome | SNOMED |
| 224960004 | Tired | SNOMED |
| 267032009 | Tired all the time | SNOMED |
| 248269005 | Tired on least exertion | SNOMED |

Mood Changes

| Code | Name | Vocabulary |
|---|---|---|
| 78667006 | Dysthymia | SNOMED |
| 274646000 | Irritability and anger | SNOMED |
| 2506003 | Early onset dysthymia | SNOMED |
| 87842000 | Generalized neuromuscular exhaustion syndrome | SNOMED |
| 19694002 | Late onset dysthymia | SNOMED |
| 724733005 | Oppositional defiant disorder co-occurrent with chronic irritability-anger | SNOMED |

| | | |
|---|---|---|
| 724734004 | Oppositional defiant disorder co-occurrent with chronic irritability-anger with normal prosocial emotions | SNOMED |
| 83176005 | Primary dysthymia | SNOMED |
| 38451003 | Primary dysthymia early onset | SNOMED |
| 67711008 | Primary dysthymia late onset | SNOMED |
| 85080004 | Secondary dysthymia | SNOMED |
| 3109008 | Secondary dysthymia early onset | SNOMED |
| 36170009 | Secondary dysthymia late onset | SNOMED |

Myalgias

| Code | Name | Vocabulary |
|---|---|---|
| 68962001 | Muscle pain | SNOMED |
| 83264000 | Epidemic pleurodynia | SNOMED |
| 95416007 | Eosinophilia myalgia syndrome | SNOMED |
| 9.57E+14 | Secondary fibromyalgia | SNOMED |
| 702549002 | Centrally mediated myalgia | SNOMED |
| 2.82E+13 | Abdominal muscle pain | SNOMED |
| 699682009 | Localized masticatory muscle soreness | SNOMED |
| 726531007 | Myofascial pain syndrome | SNOMED |
| 712752004 | Myalgia of pelvic floor | SNOMED |
| 1.65E+16 | Myalgia caused by statin | SNOMED |
| 1.14E+14 | Myofascial pain syndrome of lumbar spine | SNOMED |
| 4.13E+13 | Myofascial pain syndrome of thoracic spine | SNOMED |
| 240044003 | Fibrositis and nodular fasciitis | SNOMED |
| 240107001 | Viral myalgia | SNOMED |
| 95415006 | Polymyalgia | SNOMED |
| 22166009 | Skeletal muscle tender | SNOMED |
| 95421005 | Intercostal myalgia | SNOMED |
| 95417003 | Primary fibromyalgia syndrome | SNOMED |
| 85879003 | Secondary fibrositis | SNOMED |
| 403735006 | Eosinophilia-myalgia syndrome from tryptophan | SNOMED |

| | | |
|---|---|---|
| 76821002 | Epidemic cervical myalgia | SNOMED |
| 54981004 | Charleyhorse | SNOMED |
| 298292009 | Pain on movement of skeletal muscle | SNOMED |
| 298289005 | Muscle tender point | SNOMED |
| 298290001 | Tenderness at muscle insertion | SNOMED |
| 298288002 | Diffuse muscle tenderness | SNOMED |
| 279030006 | Myofascial pain syndrome of neck | SNOMED |
| 279070004 | Muscle tension pain | SNOMED |
| 30173002 | Primary fibrositis | SNOMED |
| 279036000 | Myofascial pain syndrome of thorax | SNOMED |
| 279041008 | Myofascial pain syndrome of lower back | SNOMED |
| 288232008 | Myalgia/myositis - ankle/foot | SNOMED |
| 288231001 | Myalgia/myositis - lower leg | SNOMED |
| 288228002 | Myalgia/myositis - forearm | SNOMED |
| 288225004 | Myalgia/myositis - multiple | SNOMED |
| 288230000 | Myalgia/myositis - pelvis/thigh | SNOMED |
| 288229005 | Myalgia/myositis - hand | SNOMED |
| 288227007 | Myalgia/myositis - upper arm | SNOMED |
| 288226003 | Myalgia/myositis - shoulder | SNOMED |
| 6.43E+15 | Muscle pain with chewing | Nebraska Lexicon |
| 8.19E+15 | Pain in the rhomboid muscle | Nebraska Lexicon |
| 3.01E+14 | Repetitive strain injury of right thigh | SNOMED |
| 3.01E+14 | Repetitive strain injury of right foot | SNOMED |
| 3.01E+14 | Repetitive strain injury of right ankle | SNOMED |
| 3.01E+14 | Repetitive strain injury of left thigh | SNOMED |
| 3.01E+14 | Repetitive strain injury of left foot | SNOMED |
| 3.01E+14 | Repetitive strain injury of left ankle | SNOMED |
| 1.57E+16 | Repetitive strain injury of bilateral forearms | SNOMED |
| 1.57E+16 | Repetitive strain injury of bilateral hands | SNOMED |

| | | |
|---|---|---|
| 56557000 | Fibrositis | SNOMED |

Myocarditis

| Code | Name | Vocabulary |
|---|---|---|
| 50920009 | Myocarditis | SNOMED |
| 46701001 | Acute myocarditis | SNOMED |
| 195033009 | Sarcoid heart muscle disease | SNOMED |
| 91025000 | Idiopathic myocarditis | SNOMED |
| 266238009 | Isolated (Fiedler's) myocarditis | SNOMED |
| 89141000 | Viral myocarditis | SNOMED |
| 22653005 | Myocarditis due to infectious agent | SNOMED |
| 194942007 | Acute myocarditis associated with another disorder | SNOMED |
| 64043005 | Bacterial myocarditis | SNOMED |
| 194750008 | Rheumatic myocarditis | SNOMED |
| 37217002 | Coxsackie myocarditis | SNOMED |
| 194709000 | Acute rheumatic myocarditis | SNOMED |
| 76534005 | Myocarditis due to acquired toxoplasmosis | SNOMED |
| 31993003 | Toxic myocarditis | SNOMED |
| 194943002 | Acute aseptic myocarditis of the newborn | SNOMED |
| 194944008 | Acute myocarditis - coxsackie | SNOMED |
| 194945009 | Acute myocarditis - diphtheritic | SNOMED |
| 194946005 | Acute myocarditis - influenzal | SNOMED |
| 194950003 | Acute myocarditis - meningococcal | SNOMED |
| 194947001 | Acute myocarditis - syphilitic | SNOMED |
| 194948006 | Acute myocarditis - toxoplasmosis | SNOMED |
| 194949003 | Acute myocarditis - tuberculous | SNOMED |
| 30328003 | Acute myoendocarditis | SNOMED |
| 11176009 | Acute myopericarditis | SNOMED |
| 65718001 | Adenoviral myocarditis | SNOMED |
| 45093008 | Chronic interstitial myocarditis | SNOMED |
| 4.51E+14 | Chronic myocarditis | SNOMED |

| | | |
|---|---|---|
| 61012002 | Chronic rheumatic heart disease with myocarditis | SNOMED |
| 756003 | Chronic rheumatic myopericarditis | SNOMED |
| 413933004 | Coxsackie myocarditis of newborn | SNOMED |
| 26117009 | Diphtheritic myocarditis | SNOMED |
| 427372006 | Eosinophilic myocarditis | SNOMED |
| 472707009 | Fetal myocarditis | SNOMED |
| 488007 | Fibroid myocarditis | SNOMED |
| 233868005 | Fungal myocarditis | SNOMED |
| 60812006 | Giant cell myocarditis | SNOMED |
| 723865005 | Infection causing myoendocarditis | SNOMED |
| 471841009 | Inflammatory cardiomyopathy | SNOMED |
| 37925008 | Interstitial myocarditis | SNOMED |
| 55482007 | Isolated diffuse granulomatous myocarditis | SNOMED |
| 427443009 | Lymphocytic myocarditis | SNOMED |
| 74918002 | Measles myocarditis | SNOMED |
| 91468009 | Meningococcal myocarditis | SNOMED |
| 63462008 | Mumps myocarditis | SNOMED |
| 421929001 | Myocarditis associated with AIDS | SNOMED |
| 1.24E+15 | Myocarditis caused by 2019 novel coronavirus | SNOMED |
| 713318009 | Myocarditis co-occurrent with human immunodeficiency virus infection | SNOMED |
| 55087008 | Myocarditis due to chemical agent | SNOMED |
| 72527006 | Myocarditis due to drug | SNOMED |
| 460345001 | Myocarditis due to echovirus | SNOMED |
| 460338001 | Myocarditis due to Genus Aspergillus | SNOMED |
| 460620006 | Myocarditis due to Genus Borrelia | SNOMED |
| 460329003 | Myocarditis due to Genus Candida | SNOMED |
| 460317007 | Myocarditis due to Genus Rickettsia | SNOMED |

| | | |
|---|---|---|
| 8676001 | Myocarditis due to hypersensitivity state | SNOMED |
| 1.06E+16 | Myocarditis due to Influenza A virus | SNOMED |
| 1.43E+14 | Myocarditis due to Influenza A virus subtype H1N1 | SNOMED |
| 78046005 | Myocarditis due to influenza virus | SNOMED |
| 460352004 | Myocarditis due to Order Spirochaetales | SNOMED |
| 79096004 | Myocarditis due to physical agent | SNOMED |
| 88782004 | Myocarditis due to radiation | SNOMED |
| 1.09E+15 | Myocarditis due to scarlet fever | SNOMED |
| 723863003 | Myopericarditis | SNOMED |
| 1.60E+16 | Myopericarditis caused by Borrelia species | SNOMED |
| 233869002 | Parasitic myocarditis | SNOMED |
| 233867000 | Q fever myocarditis | SNOMED |
| 28880005 | Rheumatoid carditis | SNOMED |
| 195136004 | Rheumatoid myocarditis | SNOMED |
| 64190005 | Rubella myocarditis | SNOMED |
| 279001 | Senile myocarditis | SNOMED |
| 194956009 | Septic myocarditis - pneumococcal | SNOMED |
| 194957000 | Septic myocarditis - staphylococcal | SNOMED |
| 194958005 | Septic myocarditis - streptococcal | SNOMED |
| 18484008 | Subacute interstitial myocarditis | SNOMED |
| 421272004 | Subacute myocarditis associated with AIDS | SNOMED |
| 69589006 | Subacute myoendocarditis | SNOMED |
| 4082005 | Syphilitic myocarditis | SNOMED |
| 187195003 | Toxoplasma myocarditis | SNOMED |
| 47292005 | Tuberculosis of myocardium | SNOMED |

SARS-CoV-2 Positive Test
SNOMED 1240580000000000 (“2019 novel coronavirus detected”)

SARS-CoV-2 Test Measurements

| Code | Name | Vocabulary |
|---|---|---|
| 94500-6 | SARS-CoV-2 (COVID-19) RNA [Presence] in Respiratory specimen by NAA with probe detection | LOINC |
| 94547-7 | SARS-CoV-2 (COVID-19) IgG+IgM Ab [Presence] in Serum or Plasma by Immunoassay | LOINC |
| 94563-4 | SARS-CoV-2 (COVID-19) IgG Ab [Presence] in Serum or Plasma by Immunoassay | LOINC |
| 94564-2 | SARS-CoV-2 (COVID-19) IgM Ab [Presence] in Serum or Plasma by Immunoassay | LOINC |
| 94762-2 | SARS-CoV-2 (COVID-19) Ab [Presence] in Serum or Plasma by Immunoassay | LOINC |
| 94558-4 | SARS-CoV-2 (COVID-19) Ag [Presence] in Respiratory specimen by Rapid immunoassay | LOINC |
| 94507-1 | SARS-CoV-2 (COVID-19) IgG Ab [Presence] in Serum, Plasma or Blood by Rapid immunoassay | LOINC |
| 94562-6 | SARS-CoV-2 (COVID-19) IgA Ab [Presence] in Serum or Plasma by Immunoassay | LOINC |
| 94508-9 | SARS-CoV-2 (COVID-19) IgM Ab [Presence] in Serum, Plasma or Blood by Rapid immunoassay | LOINC |
| 94505-5 | SARS-CoV-2 (COVID-19) IgG Ab [Units/volume] in Serum or Plasma by Immunoassay | LOINC |
| 94506-3 | SARS-CoV-2 (COVID-19) IgM Ab [Units/volume] in Serum or Plasma by Immunoassay | LOINC |
| U0002 | 2019-ncov coronavirus, sars-cov-2/2019-ncov (covid-19), any technique, multiple types | HCPCS |

| | | |
|---|---|---|
| | or subtypes (includes all targets), non-cdc | |
| 0224U | Antibody, severe acute respiratory syndrome coronavirus 2 (SARS-CoV-2) (Coronavirus disease [COVID-19]), includes titer(s), when performed | CPT4 |
| U0001 | Cdc 2019 novel coronavirus (2019-ncov) real-time rt-pcr diagnostic panel | HCPCS |
| 1.24E+15 | Detection of 2019 novel coronavirus using polymerase chain reaction technique | SNOMED |
| 86328 | Immunoassay for infectious agent antibody(ies), qualitative or semiquantitative, single step method (eg, reagent strip); severe acute respiratory syndrome coronavirus 2 (SARS-CoV-2) (Coronavirus disease [COVID-19]) | CPT4 |
| 87635 | Infectious agent detection by nucleic acid (DNA or RNA); severe acute respiratory syndrome coronavirus 2 (SARS-CoV-2) (Coronavirus disease [COVID-19]), amplified probe technique | CPT4 |
| 0202U | Infectious disease (bacterial or viral respiratory tract infection), pathogen-specific nucleic acid (DNA or RNA), 22 targets including severe acute respiratory syndrome coronavirus 2 (SARS-CoV-2), qualitative RT-PCR, nasopharyngeal swab | CPT4 |
| 0223U | Infectious disease (bacterial or viral respiratory tract infection), pathogen-specific | CPT4 |

| | | |
|---|---|---|
| | nucleic acid (DNA or RNA), 22 targets including severe acute respiratory syndrome coronavirus 2 (SARS-CoV-2), qualitative RT-PCR, nasopharyngeal swab | |
| 95380-2 | Influenza virus A and B and SARS-CoV-2 (COVID-19) and SARS-related CoV RNA panel - Respiratory specimen by NAA with probe detection | LOINC |
| 95423-0 | Influenza virus A and B and SARS-CoV-2 (COVID-19) identified in Respiratory specimen by NAA with probe detection | LOINC |
| 95422-2 | Influenza virus A and B RNA and SARS-CoV-2 (COVID-19) N gene panel - Respiratory specimen by NAA with probe detection | LOINC |
| 1.24E+15 | Measurement of 2019 novel coronavirus antibody | SNOMED |
| 1.24E+15 | Measurement of 2019 novel coronavirus antigen | SNOMED |
| OMOP4873969 | Measurement of Severe acute respiratory syndrome coronavirus 2 (SARS-CoV-2) | OMOP Extension |
| OMOP4912985 | Measurement of Severe acute respiratory syndrome coronavirus 2 (SARS-CoV-2) Genetic material using Molecular method | OMOP Extension |
| OMOP4912972 | Measurement of Severe acute respiratory syndrome coronavirus 2 (SARS-CoV-2) in Blood | OMOP Extension |
| OMOP4873967 | Measurement of Severe acute respiratory syndrome coronavirus 2 (SARS-CoV-2) in Respiratory specimen | OMOP Extension |
| OMOP4912984 | Measurement of Severe acute respiratory syndrome | OMOP Extension |

| | | |
|---|---|---|
| | coronavirus 2 (SARS-CoV-2) in Saliva | |
| OMOP4912993 | Measurement of Severe acute respiratory syndrome coronavirus 2 (SARS-CoV-2) in Sample from nose | OMOP Extension |
| OMOP4912986 | Measurement of Severe acute respiratory syndrome coronavirus 2 (SARS-CoV-2) in Specified specimen | OMOP Extension |
| OMOP4873966 | Measurement of Severe acute respiratory syndrome coronavirus 2 (SARS-CoV-2) in Unspecified specimen | OMOP Extension |
| OMOP4912974 | Measurement of Severe acute respiratory syndrome coronavirus 2 (SARS-CoV-2) using Culture method | OMOP Extension |
| OMOP4912982 | Measurement of Severe acute respiratory syndrome coronavirus 2 (SARS-CoV-2) using Nucleic acid amplification technique | OMOP Extension |
| OMOP4912973 | Measurement of Severe acute respiratory syndrome coronavirus 2 (SARS-CoV-2) using Nucleic acid amplification technique in Blood | OMOP Extension |
| OMOP4873965 | Measurement of Severe acute respiratory syndrome coronavirus 2 (SARS-CoV-2) using Nucleic acid amplification technique in Respiratory specimen | OMOP Extension |
| OMOP4912983 | Measurement of Severe acute respiratory syndrome coronavirus 2 (SARS-CoV-2) using Nucleic acid amplification technique in Saliva | OMOP Extension |
| OMOP4912992 | Measurement of Severe acute respiratory syndrome | OMOP Extension |

| | | |
|---|---|---|
| | coronavirus 2 (SARS-CoV-2) using Nucleic acid amplification technique in Sample from nose | |
| OMOP4873968 | Measurement of Severe acute respiratory syndrome coronavirus 2 (SARS-CoV-2) using Nucleic acid amplification technique in Unspecified specimen | OMOP Extension |
| OMOP4912975 | Measurement of Severe acute respiratory syndrome coronavirus 2 (SARS-CoV-2) using Sequencing | OMOP Extension |
| 94763-0 | SARS-CoV-2 (COVID-19) [Presence] in Unspecified specimen by Organism specific culture | LOINC |
| 94661-6 | SARS-CoV-2 (COVID-19) Ab [Interpretation] in Serum or Plasma | LOINC |
| 94769-7 | SARS-CoV-2 (COVID-19) Ab [Units/volume] in Serum or Plasma by Immunoassay | LOINC |
| 94504-8 | SARS-CoV-2 (COVID-19) Ab panel - Serum or Plasma by Immunoassay | LOINC |
| 94503-0 | SARS-CoV-2 (COVID-19) Ab panel - Serum, Plasma or Blood by Rapid immunoassay | LOINC |
| 94768-9 | SARS-CoV-2 (COVID-19) IgA Ab [Presence] in Serum, Plasma or Blood by Rapid immunoassay | LOINC |
| 94720-0 | SARS-CoV-2 (COVID-19) IgA Ab [Units/volume] in Serum or Plasma by Immunoassay | LOINC |
| 95125-1 | SARS-CoV-2 (COVID-19) IgA+IgM [Presence] in Serum or Plasma by Immunoassay | LOINC |
| 94761-4 | SARS-CoV-2 (COVID-19) IgG Ab [Presence] in DBS by Immunoassay | LOINC |

| | | |
|---|---|---|
| 95416-4 | SARS-CoV-2 (COVID-19) IgM Ab [Presence] in DBS by Immunoassay | LOINC |
| 94510-5 | SARS-CoV-2 (COVID-19) N gene [Cycle Threshold #] in Unspecified specimen by NAA with probe detection | LOINC |
| 94311-8 | SARS-CoV-2 (COVID-19) N gene [Cycle Threshold #] in Unspecified specimen by Nucleic acid amplification using CDC primer-probe set N1 | LOINC |
| 94312-6 | SARS-CoV-2 (COVID-19) N gene [Cycle Threshold #] in Unspecified specimen by Nucleic acid amplification using CDC primer-probe set N2 | LOINC |
| 94760-6 | SARS-CoV-2 (COVID-19) N gene [Presence] in Nasopharynx by NAA with probe detection | LOINC |
| 95409-9 | SARS-CoV-2 (COVID-19) N gene [Presence] in Nose by NAA with probe detection | LOINC |
| 94533-7 | SARS-CoV-2 (COVID-19) N gene [Presence] in Respiratory specimen by NAA with probe detection | LOINC |
| 94756-4 | SARS-CoV-2 (COVID-19) N gene [Presence] in Respiratory specimen by Nucleic acid amplification using CDC primer-probe set N1 | LOINC |
| 94757-2 | SARS-CoV-2 (COVID-19) N gene [Presence] in Respiratory specimen by Nucleic acid amplification using CDC primer-probe set N2 | LOINC |

| | | |
|---|---|---|
| 95425-5 | SARS-CoV-2 (COVID-19) N gene [Presence] in Saliva (oral fluid) by NAA with probe detection | LOINC |
| 94766-3 | SARS-CoV-2 (COVID-19) N gene [Presence] in Serum or Plasma by NAA with probe detection | LOINC |
| 94316-7 | SARS-CoV-2 (COVID-19) N gene [Presence] in Unspecified specimen by NAA with probe detection | LOINC |
| 94307-6 | SARS-CoV-2 (COVID-19) N gene [Presence] in Unspecified specimen by Nucleic acid amplification using CDC primer-probe set N1 | LOINC |
| 94308-4 | SARS-CoV-2 (COVID-19) N gene [Presence] in Unspecified specimen by Nucleic acid amplification using CDC primer-probe set N2 | LOINC |
| 95411-5 | SARS-CoV-2 (COVID-19) neutralizing antibody [Presence] in Serum by pVNT | LOINC |
| 95410-7 | SARS-CoV-2 (COVID-19) neutralizing antibody [Titer] in Serum by pVNT | LOINC |
| 94644-2 | SARS-CoV-2 (COVID-19) ORF1ab region [Cycle Threshold #] in Respiratory specimen by NAA with probe detection | LOINC |
| 94511-3 | SARS-CoV-2 (COVID-19) ORF1ab region [Cycle Threshold #] in Unspecified specimen by NAA with probe detection | LOINC |
| 94559-2 | SARS-CoV-2 (COVID-19) ORF1ab region [Presence] in | LOINC |

| | | |
|---|---|---|
| | Respiratory specimen by NAA with probe detection | |
| 94639-2 | SARS-CoV-2 (COVID-19) ORF1ab region [Presence] in Unspecified specimen by NAA with probe detection | LOINC |
| 94646-7 | SARS-CoV-2 (COVID-19) RdRp gene [Cycle Threshold #] in Respiratory specimen by NAA with probe detection | LOINC |
| 94645-9 | SARS-CoV-2 (COVID-19) RdRp gene [Cycle Threshold #] in Unspecified specimen by NAA with probe detection | LOINC |
| 94534-5 | SARS-CoV-2 (COVID-19) RdRp gene [Presence] in Respiratory specimen by NAA with probe detection | LOINC |
| 94314-2 | SARS-CoV-2 (COVID-19) RdRp gene [Presence] in Unspecified specimen by NAA with probe detection | LOINC |
| 94745-7 | SARS-CoV-2 (COVID-19) RNA [Cycle Threshold #] in Respiratory specimen by NAA with probe detection | LOINC |
| 94746-5 | SARS-CoV-2 (COVID-19) RNA [Cycle Threshold #] in Unspecified specimen by NAA with probe detection | LOINC |
| 94819-0 | SARS-CoV-2 (COVID-19) RNA [Log #/volume] (viral load) in Unspecified specimen by NAA with probe detection | LOINC |
| 94565-9 | SARS-CoV-2 (COVID-19) RNA [Presence] in Nasopharynx by NAA with non-probe detection | LOINC |
| 94759-8 | SARS-CoV-2 (COVID-19) RNA [Presence] in Nasopharynx by NAA with probe detection | LOINC |

| | | |
|---|---|---|
| 95406-5 | SARS-CoV-2 (COVID-19) RNA [Presence] in Nose by NAA with probe detection | LOINC |
| 95424-8 | SARS-CoV-2 (COVID-19) RNA [Presence] in Respiratory specimen by Sequencing | LOINC |
| 94845-5 | SARS-CoV-2 (COVID-19) RNA [Presence] in Saliva (oral fluid) by NAA with probe detection | LOINC |
| 94822-4 | SARS-CoV-2 (COVID-19) RNA [Presence] in Saliva (oral fluid) by Sequencing | LOINC |
| 94660-8 | SARS-CoV-2 (COVID-19) RNA [Presence] in Serum or Plasma by NAA with probe detection | LOINC |
| 94309-2 | SARS-CoV-2 (COVID-19) RNA [Presence] in Unspecified specimen by NAA with probe detection | LOINC |
| 94531-1 | SARS-CoV-2 (COVID-19) RNA panel - Respiratory specimen by NAA with probe detection | LOINC |
| 94306-8 | SARS-CoV-2 (COVID-19) RNA panel - Unspecified specimen by NAA with probe detection | LOINC |
| 94642-6 | SARS-CoV-2 (COVID-19) S gene [Cycle Threshold #] in Respiratory specimen by NAA with probe detection | LOINC |
| 94643-4 | SARS-CoV-2 (COVID-19) S gene [Cycle Threshold #] in Unspecified specimen by NAA with probe detection | LOINC |
| 94640-0 | SARS-CoV-2 (COVID-19) S gene [Presence] in Respiratory specimen by NAA with probe detection | LOINC |
| 94767-1 | SARS-CoV-2 (COVID-19) S gene [Presence] in Serum or Plasma by NAA with probe detection | LOINC |

| | | |
|---|---|---|
| 94641-8 | SARS-CoV-2 (COVID-19) S gene [Presence] in Unspecified specimen by NAA with probe detection | LOINC |
| 94764-8 | SARS-CoV-2 (COVID-19) whole genome [Nucleotide sequence] in Isolate by Sequencing | LOINC |
| 94313-4 | SARS-like coronavirus N gene [Cycle Threshold #] in Unspecified specimen by NAA with probe detection | LOINC |
| 94310-0 | SARS-like coronavirus N gene [Presence] in Unspecified specimen by NAA with probe detection | LOINC |
| 94509-7 | SARS-related coronavirus E gene [Cycle Threshold #] in Unspecified specimen by NAA with probe detection | LOINC |
| 94758-0 | SARS-related coronavirus E gene [Presence] in Respiratory specimen by NAA with probe detection | LOINC |
| 94765-5 | SARS-related coronavirus E gene [Presence] in Serum or Plasma by NAA with probe detection | LOINC |
| 94315-9 | SARS-related coronavirus E gene [Presence] in Unspecified specimen by NAA with probe detection | LOINC |
| 94502-2 | SARS-related coronavirus RNA [Presence] in Respiratory specimen by NAA with probe detection | LOINC |
| 94647-5 | SARS-related coronavirus RNA [Presence] in Unspecified specimen by NAA with probe detection | LOINC |
| 94532-9 | SARS-related coronavirus+MERS coronavirus RNA [Presence] | LOINC |

| | |
|---|---|
| | in Respiratory specimen by NAA with probe detection |

Sleep Disorder

| Code | Name | Vocabulary |
|---|---|---|
| 193462001 | Insomnia | SNOMED |
| 39898005 | Sleep disorder | SNOMED |
| 77692006 | Hypersomnia | SNOMED |
| 44186003 | Dyssomnia | SNOMED |
| 3972004 | Primary insomnia | SNOMED |
| 425832009 | Psychophysiologic insomnia | SNOMED |
| 248256006 | Not getting enough sleep | SNOMED |
| 9.15E+13 | Narcolepsy without cataplexy | SNOMED |
| 194437008 | Disorders of initiating and maintaining sleep | SNOMED |
| 81608000 | Insomnia disorder related to known organic factor | SNOMED |
| 415238003 | REM sleep behavior disorder | SNOMED |
| 271794005 | Disorder of sleep-wake cycle | SNOMED |
| 80623000 | Sleep-wake schedule disorder, delayed phase type | SNOMED |
| 193042000 | Cataplexy and narcolepsy | SNOMED |
| 24121004 | Insomnia disorder related to another mental disorder | SNOMED |
| 472819006 | Adjustment insomnia | SNOMED |
| 441910000 | Idiopathic sleep related non-obstructive alveolar hypoventilation | SNOMED |
| 270487001 | Non-organic sleep disorder | SNOMED |
| 58690002 | Parasomnia | SNOMED |
| 442416002 | Idiopathic hypersomnia associated with long sleep time | SNOMED |
| 442176004 | Sleep dysfunction with arousal disturbance | SNOMED |
| 399040002 | Congenital central hypoventilation | SNOMED |
| 8.99E+13 | Sleep related hypoventilation or hypoxemia | SNOMED |
| 89675003 | Sleep terror disorder | SNOMED |

| | | |
|---|---|---|
| 80495009 | Sleep walking disorder | SNOMED |
| 418475009 | Drug-induced sleep disorder | SNOMED |
| 713498009 | Circadian rhythm sleep disorder of shift work type | SNOMED |
| 443760008 | Sleep hypoventilation | SNOMED |
| 430893009 | Sleep related rhythmic movement disorder | SNOMED |
| 9.04E+13 | Behavioral insomnia of childhood | SNOMED |
| 268722008 | Non-organic disorder of the sleep-wake schedule | SNOMED |
| 274950005 | Sleep related bruxism | SNOMED |
| 271793004 | Irregular sleep-wake pattern | SNOMED |
| 442292004 | Idiopathic hypersomnia without long sleep time | SNOMED |
| 2.80E+13 | Insomnia co-occurrent and due to medical condition | SNOMED |
| 192454004 | Nonorganic insomnia | SNOMED |
| 31537005 | Sleep-wake schedule disorder, advanced phase type | SNOMED |
| 41083005 | Alcohol-induced sleep disorder | SNOMED |
| 441830002 | Confusional arousal disorder | SNOMED |
| 426451004 | Recurrent hypersomnia | SNOMED |
| 442120007 | Recurrent isolated sleep paralysis | SNOMED |
| 2.88E+14 | Behavioral insomnia of childhood, sleep onset association type | SNOMED |
| 441976002 | Organic parasomnia | SNOMED |
| 268654005 | Repetitive intrusions of sleep | SNOMED |
| 230496009 | Non-24 hour sleep-wake cycle | SNOMED |
| 2.88E+14 | Behavioral insomnia of childhood, limit setting type | SNOMED |
| 89415002 | Hypersomnia disorder related to another mental disorder | SNOMED |
| 9.00E+13 | Organic sleep related movement disorder | SNOMED |
| 36124002 | Primary hypersomnia | SNOMED |

| | | |
|---|---|---|
| 44455001 | Hypersomnia disorder related to a known organic factor | SNOMED |
| 2.88E+14 | Behavioral insomnia of childhood, combined type | SNOMED |
| 418955009 | Organic sleep disorder | SNOMED |
| 57588009 | Sedative, hypnotic AND/OR anxiolytic-induced sleep disorder | SNOMED |
| 88926005 | Opioid-induced sleep disorder | SNOMED |
| 22574000 | Cocaine-induced sleep disorder | SNOMED |
| 724751006 | Sleep-wake schedule disorder due to jet lag | SNOMED |
| 1.05E+14 | Acquired central alveolar hypoventilation | SNOMED |
| 762348004 | Acute insomnia | SNOMED |
| 248262001 | Always sleepy | SNOMED |
| 25753007 | Amphetamine-induced sleep disorder | SNOMED |
| 787175002 | ANK3-related intellectual disability, sleep disturbance syndrome | SNOMED |
| 722293005 | Autosomal dominant cerebellar ataxia, deafness and narcolepsy syndrome | SNOMED |
| 724749007 | Behaviorally induced hypersomnia | SNOMED |
| 413638006 | Benign neonatal sleep myoclonus | SNOMED |
| 111489007 | Breathing-related sleep disorder | SNOMED |
| 56194001 | Caffeine-induced sleep disorder | SNOMED |
| 24825006 | Central alveolar hypoventilation syndrome | SNOMED |
| 724748004 | Chronic insomnia | SNOMED |
| 9.15E+13 | Circadian rhythm disorder caused by drug | SNOMED |
| 1.35E+14 | Circadian rhythm sleep disorder caused by alcohol | SNOMED |

| | | |
|---|---|---|
| 248258007 | Circumstances interfere with sleep | SNOMED |
| 3.18E+13 | Daytime hypersomnia | SNOMED |
| 194439006 | Disorders of excessive somnolence | SNOMED |
| 3.15E+13 | Disruptions of 24 hour sleep-wake cycle | SNOMED |
| 711565008 | Dream enactment behavior | SNOMED |
| 9.15E+13 | Drug-induced hypersomnia | SNOMED |
| 401236004 | Early morning waking | SNOMED |
| 429276004 | Environmental sleep disorder | SNOMED |
| 230490003 | Excessive day and night-time sleepiness | SNOMED |
| 230489007 | Excessive daytime sleepiness - normal night sleep | SNOMED |
| 230492006 | Excessive daytime sleepiness with sleep paralysis | SNOMED |
| 449160007 | Exploding head syndrome | SNOMED |
| 715829003 | Familial advanced sleep phase syndrome | SNOMED |
| 83157008 | Fatal familial insomnia | SNOMED |
| 719972004 | Haddad syndrome | SNOMED |
| 1.35E+14 | Hypersomnia caused by alcohol | SNOMED |
| 426943005 | Hypersomnia disorder related to menstruation | SNOMED |
| 230488004 | Hypersomnia of non-organic origin | SNOMED |
| 724750007 | Hypersomnolence caused by substance | SNOMED |
| 724752004 | Hypnogogic exploding head syndrome | SNOMED |
| 2.88E+14 | Hyposomnia co-occurrent and due to psychological disorder | SNOMED |
| 429456008 | Hypoventilation during sleep due to neuromuscular disorder | SNOMED |
| 3.73E+12 | Idiopathic hypersomnia | SNOMED |
| 2.35E+16 | Idiopathic hypersomnolence | Nebraska Lexicon |
| 59050008 | Initial insomnia | SNOMED |
| 9.37E+13 | Insomnia co-occurrent and due to nocturnal myoclonus | SNOMED |

| | | |
|---|---|---|
| 4.36E+14 | Insomnia due to anxiety and fear | SNOMED |
| 111488004 | Kleine-Levin syndrome | SNOMED |
| 162204000 | Late insomnia | SNOMED |
| 724508005 | Late-onset central hypoventilation co-occurrent and due to hypothalamic dysfunction | SNOMED |
| 425881006 | Limit-setting sleep disorder | SNOMED |
| 443664006 | Long sleeper syndrome | SNOMED |
| 198437004 | Menopausal sleeplessness | SNOMED |
| 67233009 | Middle insomnia | SNOMED |
| 54230003 | Mixed insomnia | SNOMED |
| 60380001 | Narcolepsy | SNOMED |
| 735676003 | Narcolepsy type 1 | SNOMED |
| 3.41E+13 | Nocturnal myoclonus | SNOMED |
| 407666007 | Nocturnal sleep-related eating disorder | SNOMED |
| 425476007 | Non-organic parasomnia | SNOMED |
| 1.06E+14 | Organic mixed sleep disorder | SNOMED |
| 9.97E+13 | Organic sleep-wake cycle disorder | SNOMED |
| 248261008 | Oversleeps | SNOMED |
| 17402007 | Pavor diurnus | SNOMED |
| 191999000 | Persistent hypersomnia | SNOMED |
| 191997003 | Persistent insomnia | SNOMED |
| 418424006 | Post-radiotherapy somnolence syndrome | SNOMED |
| 230491004 | Postviral excessive daytime sleepiness | SNOMED |
| 789009003 | Primary central sleep apnea of prematurity | SNOMED |
| 1.05E+14 | Primary hyposomnia | SNOMED |
| 247951006 | Reaction to sudden wakening | SNOMED |
| 88982005 | Rebound insomnia | SNOMED |
| 724753009 | Recurrent isolated sleep-related hallucinations | SNOMED |
| 192004002 | Repeated rapid eye movement sleep interruptions | SNOMED |
| 192008004 | Reversed sleep-wake cycle | SNOMED |
| 4.34E+14 | Secondary narcolepsy | SNOMED |

| | | |
|---|---|---|
| 4.34E+14 | Secondary narcolepsy with cataplexy | SNOMED |
| 4.34E+14 | Secondary narcolepsy without cataplexy | SNOMED |
| 428708005 | Secondary parasomnia | SNOMED |
| 427426006 | Sleep attack | SNOMED |
| 247962006 | Sleep automatism | SNOMED |
| 723937009 | Sleep disorder caused by cannabis | SNOMED |
| 1.24E+16 | Sleep disorder caused by methamphetamine | SNOMED |
| 724699001 | Sleep disorder caused by nicotine | SNOMED |
| 724732000 | Sleep disorder caused by psychoactive substance | SNOMED |
| 713524002 | Sleep disorder caused by reverse transcriptase inhibitor | SNOMED |
| 762334004 | Sleep disorder caused by stimulant | SNOMED |
| 737342004 | Sleep disorder caused by synthetic cannabinoid | SNOMED |
| 762518003 | Sleep disorder caused by synthetic cathinone | SNOMED |
| 4.68E+12 | Sleep disturbance in infancy | SNOMED |
| 192003008 | Sleep drunkenness | SNOMED |
| 441877007 | Sleep dysfunction with sleep stage disturbance | SNOMED |
| 426542005 | Sleep hypoventilation due to lower airway obstruction | SNOMED |
| 277180005 | Sleep paralysis | SNOMED |
| 2.89E+14 | Sleep related hypoxemia | SNOMED |
| 8.50E+13 | Sleep related movement disorder | SNOMED |
| 277181009 | Sleep starts | SNOMED |
| 426824009 | Sleep-onset association disorder | SNOMED |
| 429571005 | Sleep-related dissociative disorder | SNOMED |
| 230500006 | Sleep-related dystonia | SNOMED |
| 429266002 | Sleep-related groaning | SNOMED |
| 230497000 | Sleep-related head banging | SNOMED |

| | | |
|---|---|---|
| 724509002 | Sleep-related hypoventilation caused by substance | SNOMED |
| 789046004 | Sleep-related movement disorder caused by drug | SNOMED |
| 789047008 | Sleep-related movement disorder caused by substance | SNOMED |
| 430390000 | Sleep-related neurogenic tachypnea | SNOMED |
| 230498005 | Sleep-related painful erections | SNOMED |
| 230499002 | Sleep-related respiratory failure | SNOMED |
| 54532007 | Sleep-wake schedule disorder, frequently changing type | SNOMED |
| 2.39E+13 | Somnambulism co-occurrent with sleep terror disorder | SNOMED |
| 370971007 | Somnolence syndrome | SNOMED |
| 363314000 | Substance-induced sleep disorder | SNOMED |
| 248259004 | Symptoms interfere with sleep | SNOMED |
| 67062000 | Terminal insomnia | SNOMED |
| 268653004 | Transient hypersomnia | SNOMED |
| 268652009 | Transient insomnia | SNOMED |
| 230495008 | Transient sleep-wake rhythm disorder | SNOMED |
| 248260009 | Unrefreshed by sleep | SNOMED |

Tachycardia

| Code | Name | Vocabulary |
|---|---|---|
| 80313002 | Palpitations | SNOMED |
| 3424008 | Tachycardia | SNOMED |
| 278086000 | Baseline tachycardia | SNOMED |
| 161968007 | Bumping heart | SNOMED |
| 462170003 | Fetal supraventricular tachycardia with long ventriculoatrial interval | SNOMED |
| 462169004 | Fetal supraventricular tachycardia with short ventriculoatrial interval | SNOMED |
| 240298005 | Fetal tachycardia | SNOMED |

| | | |
|---|---|---|
| 4006006 | Fetal tachycardia affecting management of mother | SNOMED |
| 161969004 | Fluttering heart | SNOMED |
| 102590007 | Intermittent palpitations | SNOMED |
| 2.40E+14 | Irregular palpitations | SNOMED |
| 162992001 | O/E - pulse rate tachycardia | SNOMED |
| 248648003 | Palpitations - rapid | SNOMED |
| 428919002 | Palpitations with regular rhythm | SNOMED |
| 709065005 | Postoperative fluttering heart | SNOMED |
| 248657009 | Pounding heart | SNOMED |
| 442515000 | Reflex tachycardia | SNOMED |
| 2.40E+14 | Regular palpitations | SNOMED |
| 11092001 | Sinus tachycardia | SNOMED |
| 421348006 | Tachycardia - baseline fetal heart rate | SNOMED |
| 74615001 | Tachycardia-bradycardia | SNOMED |

SNOMED, Systematic Nomenclature of Medicine; ICD9CM, International Classification of Diseases Ninth Revision Clinical Modification; HCPCS, Healthcare Common Procedure Coding System; CPT4, Current Procedural Terminology 4;OMOP, Observational Medical Outcomes Partnership
