## Supplementary Appendix#3 for "Characterizing Post-Acute Sequelae of SARS-CoV-2 Infection across Claims and Electronic Health Record Databases"

Supplementary Appendix#3: Calculation of Relative Risk

*Calculation of Relative Risk:*
To attain a point estimate of relative risk for PASC, COVID-19 relative to influenza patients, we divided the proportion of patients with any of the PASC related symptoms or diagnoses in the COIVD-19 cohort by the corresponding proportion in the influenza cohort. We used that measure of association to compare relative risk across databases. For our average relative risk, the CUIMC database had one tenth the weight of the other databases due to differences in sample size. Additionally, we calculated the relative risk for each individual PASC related diagnosis or symptom.
